# Trends in the relationship between psychological distress and depression diagnosis in the general adult population 2011-2022

**DOI:** 10.64898/2026.08.17.26360443

**Authors:** Thomas Steare, Sally McManus, Matthias Pierce, Praveetha Patalay

## Abstract

**Background:** Various explanations have been proposed for increasing trends in diagnosed depression in the UK, including increases in the proportion of the population that experience symptoms, changes in the threshold for seeking treatment and changes in clinical recognition or coding practices. Identifying trends over time for the relationship between the experiences of psychological distress and receiving a diagnosis can help explain wider trends in the incidence of clinical depression, such as whether the threshold for seeking treatment and receiving a diagnosis of depression has changed.

**Aims:** This study aims to examine trends in the incidence of diagnosed depression, and relationships between psychological distress and recent depression diagnosis among UK adults between 2011 and 2022. We also assess whether the difference in psychological distress between adults with and without a recent depression diagnosis has changed over time and examine these relationships across subgroups (sex, ethnicity, age, cohort, education and financial stress).

**Methods:** Data were from 66,360 adults (341,764 observations) aged 16 or older from the UK Household Longitudinal Study (UKHLS) across nine fieldwork periods spanning 2011-2022. Psychological distress was reported with the GHQ-12 used as a continuous variable and as a binary variable indicating caseness. Recent depression diagnoses were self-reported. Analyses we run for the overall population and stratified by different sociodemographic characteristics.

**Results:** Incidence of diagnosed depression has not increased over time in the overall sample, but there was a notable increase in some sub-groups, most clearly seen for women aged 16 to 24. There has been a clear increase in the number of cases of psychological distress, but who have not received a recent diagnosis of depression. The level of psychological distress experienced by adults recently diagnosed with depression has slightly increased over time, whilst the difference in psychological distress experienced by adults with and without a recent depression diagnosis remained stable. Subgroup analyses show differences in the distress experienced by those with and without a recent diagnosis based on sex, age, cohort, ethnicity, education and financial situation: temporal trends were mostly similar across groups.

**Conclusions:** Stable trends in (a) the distress experienced by adults recently diagnosed with depression, and (b) the difference in psychological distress experienced by adults with a recent depression diagnosis compared to adults without suggests little support for the hypothesis that depression is being diagnosed at lower levels of psychological distress. Instead, our findings suggest there may be a growing population who are not receiving clinical support for high levels of distress.

## Introduction

Over the past 30 years in the UK, there has been a trend of worsening mental health in the adult population.^1,2^ This trend is evident across different measurements and study designs, including in surveys that have found an increase in self-reported symptoms of psychological distress, and in healthcare data that has shown an increase in the incidence of common mental disorder, such as depression, and a rise in the number of people receiving mental health treatment.^1–4^ Explanations for increases in diagnosed common mental disorder include worsening distress levels in the population, changes in sub-populations seeking healthcare, a lowering of the distress threshold that people seek mental healthcare, and changes in diagnostician practices, such as a potential lowering of thresholds where a diagnosis is given.^5^

Trends in self-reported distress are typically estimated using population-based surveys, whilst trends in healthcare access and diagnoses are typically estimated using electronic health records for the latter, with little research able to study the relationships between symptoms and diagnosis in the same people and over time. Moreover, we know that experience of symptoms, healthcare access and receiving a diagnosis are unequally distributed by sociodemographic and economic factors such as sex, age, ethnicity and socioeconomic resources.^6,7^ It is also possible that the relationships between symptoms and receiving diagnoses are changing over time differently for different subgroups.

We present trends across approximately a decade in (a) the incidence of newly diagnosed depression, (b) the level of psychological distress in adults with a recent diagnosis of depression, and (c) the proportion of adults that report high psychological distress but do not have a recent depression diagnosis. We also estimate the difference in psychological distress between adults with a recent depression diagnosis and adults who have never been diagnosed at each timepoint and plot whether this has changed over time. We conducted additional stratified analyses by sex, age, cohort, ethnicity, education-level, and financial stress to examine whether there are subgroup differences in temporal trends.

## Methods

### Sample

We used data from a nationally representative household panel study, the UK Household Longitudinal Study (UKHLS), also known as ‘Understanding Society’.^8^ As of August 2026, data is available from fifteen consecutive annual waves.

During the initial recruitment period (2009-2011), approximately 26,000 households were recruited through (a) a clustered and stratified probability sample of households across Great Britain, and (b) a simple random sample of households in Northern Ireland. At this time, an Ethnic Minority Boost sample of approximately 4,000 households was also added. A further Immigrant and Ethnicity Boost sample of approximately 2,500 households was added in 2015, with 5,800 further households added in 2022-2023 as part of a General Population Sample boost. At each wave, the UKHLS adult sample consists of returning participants who have participated in at least one previous wave and new entrants (e.g., adults from newly recruited households, participants turning 16 and eligible for their first adult survey), who are taking part in their first wave, apart from the first wave where all participants are new entrants.

We included data from nine waves that we deemed to measure depression diagnoses in a sufficiently comparable way. In these waves, most participants were returning participants who self-report new diagnoses since the last wave, with a smaller proportion of the sample being new entrants who report lifetime history, from which new past-year diagnosis could be derived.. We used a complete case sample, where the sample at each timepoint consisted of those with available data for all variables of interest specific to each analysis.

We used data from participants aged 16 or over who completed the adult survey in any of the nine timepoints of our study (Table 1). We use the term timepoint to refer to the fieldwork periods in our study, contrasting to wave which we use when referring to the timepoints in the UKHLS (Table 1). The first timepoint was carried out between January 2011 to April 2013, and the ninth and final timepoint was carried out between January 2020 to May 2022.

**Table 1.** Timepoints within our study.

| Study timepoint | UKHLS Wave | Fieldwork period | Reason for exclusion |
| --- | --- | --- | --- |
|  | 1 | January 2009 – March 2011 | All participants are new entrants and are therefore asked only about lifetime history of health conditions. Wave 1 therefore contrasts to preceding waves where most of the sample are returning members who report health conditions diagnosed since the previous wave. |
|  | 2 | January 2010 – March 2012 | Only returning participants are asked about health conditions, therefore new entrants would be excluded from this wave. |
| 1 | 3 | January 2011 – April 2013 |  |
| 2 | 4 | January 2012 – May 2014 |  |
| 3 | 5 | January 2013 – April 2015 |  |
| 4 | 6 | January 2014 – May 2016 |  |
| 5 | 7 | January 2015 – May 2017 |  |
| 6 | 8 | January 2016 – May 2018 |  |
| 7 | 9 | January 2017 – May 2019 |  |
|  | 10 | January 2018 – May 2020 | Returning participants are not asked about new clinical conditions since the previous wave and instead are asked about lifetime history. |
| 8 | 11 | January 2019 – May 2021 |  |
| 9 | 12 | January 2020 – May 2022 |  |
|  | 13 | January 2021 – May 2023 | For the first time, mental and physical health conditions are assessed separately. This could potentially affect reporting due to differences in how these questions are framed compared to in earlier waves. |
|  | 14 | January 2022 – May 2024 |  |
|  | 15 | January 2023 – May 2025 |  |

### Recent depression diagnosis

We used self-reported diagnosis of depression and focussed on recent diagnosis to ensure consistency across the sample. At all timepoints, adults new to the survey are asked to report on any diagnoses given by a health professional in their lifetime. Participants are further asked about the age they developed the condition and whether symptoms of this condition are still present. Participants that have previously completed a survey are asked whether they have received any new clinical diagnoses since they last completed an adult survey in the UKHLS. Participants are not prohibited from reporting a new diagnosis of depression if they have previously reported a diagnosis in preceding waves.

From waves 1 to 9 of the UKHLS, adult participants are presented with a list of health conditions, with “clinical depression” one of the conditions listed. From wave 10 to 12 “clinical depression” is no longer an option and is replaced with an “emotional, nervous or psychiatric problem”. Participants that report an emotional, nervous or psychiatric problem are then asked what diagnosis they have, with “depression” one of the response options alongside other mental health problems. From wave 13, mental and physical health problems are assessed separately. Participants are asked which mental health conditions if any, they have been diagnosed with by a health professional. Item wording is presented in Table 2.

**Table 2.**
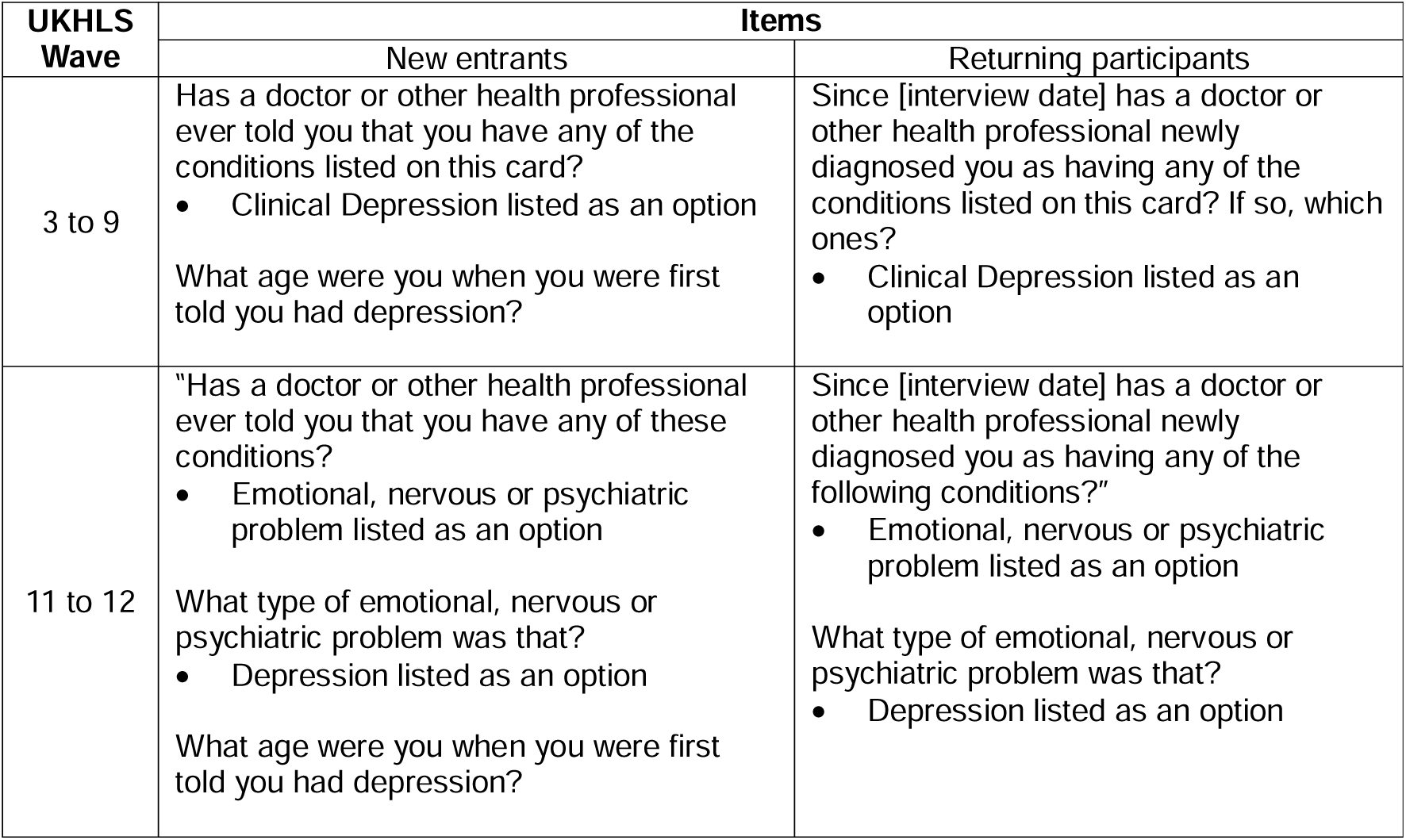
Items used to assess whether new entrants have ever received a clinical diagnosis of depression.

| UKHLS Wave | Items |  |
| --- | --- | --- |
|  | New entrants | Returning participants |
| 3 to 9 | Has a doctor or other health professional ever told you that you have any of the conditions listed on this card?<br>Clinical Depression listed as an option<br>What age were you when you were first told you had depression? | Since [interview date] has a doctor or other health professional newly diagnosed you as having any of the conditions listed on this card? If so, which ones?<br>Clinical Depression listed as an option |
| 11 to 12 | “Has a doctor or other health professional ever told you that you have any of these conditions?<br>Emotional, nervous or psychiatric problem listed as an option<br>What type of emotional, nervous or psychiatric problem was that?<br>Depression listed as an option<br>What age were you when you were first told you had depression? | Since [interview date] has a doctor or other health professional newly diagnosed you as having any of the following conditions?”<br>Emotional, nervous or psychiatric problem listed as an option<br>What type of emotional, nervous or psychiatric problem was that?<br>Depression listed as an option |

For new entrants, who were asked about lifetime history of diagnosed depression, we classed a new case of diagnosed depression as an adult who reported a diagnosis that was first made no more than one year ago, to closely align to the intervals between annual UKHLS waves. This was calculated through identifying the difference between the participants’ age at the current timepoint and the self-reported age at first diagnosis. Participants’ age and the age they report when they were diagnosed are reported in whole years. In extreme cases, the difference between them could be reported as a single year, when in fact the true value is closer to two years. As a result, a number of new entrants might inadvertently be classed as new cases of diagnosed depression.

For returning participants, an adult who stated they have been newly diagnosed with depression since the last wave they attended were classed as a new case. In many cases, this period represents a year but will be longer if participants did not attend the previous wave.

### Psychological distress

At each timepoint, participants self-complete the 12-item General Health Questionnaire (GHQ-12).^9^ For each item, participants indicate on a four-point scale the extent to which they have felt a certain way compared to usual. The GHQ-12 is a validated and widely used measure of psychological distress. It is not specifically designed to measure depressive symptoms; however, it features many items that assess symptoms and experiences that are central to depression such as low mood, a limited ability to enjoy activities and disrupted sleep.

The GHQ-12 can be scored in two ways. Bimodal scoring involves applying a score of 1 for each item where the participant indicates one of the two options representing greater psychological distress, and a score of 0 for options less indicative of psychological distress, leading to potential score range of 0-12. For the Likert scoring approach, responses are scaled to 0 to 3, with a scores range of 0-36. For both approaches, a greater score indicates greater levels of psychological distress.

For our analyses we use (a) a caseness approach of psychological distress, with scores of 4 or greater on the bimodal approach classed as meeting caseness for psychological distress,^10,11^ and (b) a continuous score derived from the Likert scoring system.

### Diagnosis and psychological distress

We created a categorical variable at each timepoint based on whether adults (a) met caseness for psychological distress and (b) had received a recent diagnosis of clinical depression. At each timepoint we estimated if adults meet psychological distress caseness and have not received a recent clinical diagnosis of depression (distressed with no recent diagnosis).

### Personal characteristics

The analyses were stratified according to the following variables.

#### Sex

Participants’ sex was recorded as male or female. When stratifying by sex, we excluded adults inconsistently classed (reported a different sex across waves) due to this applying to a very small proportion of the sample (<0.01%), meaning any estimates would be extremely imprecise.

#### Age

We categorised age into four bandings: 16 to 24, 25 to 44, 45 to 64, and 65 or older. Use of these groupings was based on considering the trade-offs of using narrow age groups at the expense of statistical power, and therefore reducing precision in the analyses, and capturing age groups that are meaningful.

#### Cohort

We created a variable regarding adults’ birth cohort based on their year of birth. Participants born before 1946 were classed as Pre-Boomers, adults born between 1946 and 1964 were classed as Baby Boomers, adults born between 1965 and 1980 were classed as Generation X and adults born between 1981 and 1996 were classed as Millennials. We did not include Generation Z (born between 1997 and 2011) in cohort-stratified analyses as only two Generation Z participants were eligible for the adult surveys at our first study timepoint, and the youngest Generation Z participants were never eligible for any adult survey during the whole study period.

#### Ethnicity

We used a binary classification of ethnicity due to sample size constrains. Adults that self-reported being White British, White Irish, gypsy or Irish Traveller or any other White background were classed as White. We grouped adults of non-White ethnicities into an ethnic minority category. We used this binary grouping as sample sizes for different ethnicities are not large enough to produce accurate and stable estimates. We also report supplementary analyses with a four-category grouping (White, Mixed, Asian, and Black), we do not estimate results for adults from other ethnicities that do not fit the four categories, due to the small size of this subgroup (<1% of all observations) and therefore low precision for any resultant estimates.

#### Education level

Adults’ highest level of education attained is self-reported by participants when they first complete an adult survey and is updated if they report any further qualifications at future waves. We used four categories: (a) higher education, including degrees and other higher qualifications, (b) upper secondary, such as A-Levels or equivalents, (c) lower secondary, such as GCSEs or equivalents, and (d) no qualifications. We did not include participants that report “other qualifications” when stratifying by education level as assignment to this group is hard to interpret.

#### Financial stress

At each UKHLS wave adults self-report their level of financial stress via a single item; “How well would you say you yourself are managing financially these days?”. We created three groups based on the possible response options: (a) adults that are “living comfortably” or are “doing all right”, (b) adults that are “just about getting by”, and (c) adults that are “finding it quite difficult” or “very difficult”.

### Data analysis

We estimated the proportion of the sample at each timepoint that reported receiving a new diagnosis of depression and plotted these estimates over the nine timepoints. We examined changes in the relationship between recent depression diagnosis and psychological distress by estimating at each timepoint; (a) the mean GHQ-12 score (Likert scoring) in adults with a recent diagnosis of depression and (b) the proportion of the sample that meet caseness of psychological distress (score of 4 or greater on the GHQ-12; bimodal scoring) but do not have a recent diagnosis of depression. We also estimated the mean difference in GHQ-12 scores between adults with a recent clinical depression diagnosis and adults without a recent diagnosis using linear regression at each timepoint. Analyses were stratified according to different personal characteristics. We ran no formal modelling to assess trends.

For all analyses, we took into account the complex survey design and applied the cross-sectional sampling weights applicable for each timepoint to account for non-response and correct for unequal selection probabilities. We did not impute missing data. As the sample at each timepoint consists of different compositions of survey members, analyses represent trends in population-level estimates rather than changes within a fixed cohort.

We dropped estimates from figures where 95% confidence intervals could not be estimated because there were insufficient design degrees of freedom. Additionally, we dropped estimates from figures where confidence intervals were extremely wide and subsequently hindered figure presentation and interpretation. These all occurred for small subgroups. We report all estimates in the supplementary material (Supplementary Tables 1 to 8).

We re-ran all analyses, where we excluded from our sample at each timepoint any participant that had reported a new diagnosis of depression at any previous timepoint in our study, as well as at the UKHLS wave 1 where lifetime history of depression is reported. This approach and the approach used in the main analyses have different strengths and different limitations that might lead to different biases (Table 3). As such we report results using both approaches.

**Table 3.** Advantages and limitations of different approaches to handling participants in future waves after reporting they have received a new diagnosis of depression.

|  | Including participants in future timepoints after reporting a new diagnosis of depression at a previous timepoint | Excluding participants from future timepoints after reporting a new diagnosis of depression |
| --- | --- | --- |
| Advantages | The sample composition stays as consistent as possible, excluding changes due to study attrition and sample boosts. | Adults can only ever report receiving a new diagnosis once, meaning estimates of new cases of depression are likely to be more accurate.<br>Excluding adults with prior depression that re-report a new diagnosis in the study, might increase the accuracy of the estimated level of psychological distress of adults with a new depression diagnosis, as it excludes adults with potentially chronic depression that could be accompanied by severe psychological distress. |
| Limitations | Adults can report receiving a new diagnosis across multiple waves, causing inflated estimates of new cases of depression at all timepoints.<br>Including adults with past depression with long-lasting psychological distress could lead to overestimating the number of adults that report psychological distress but do not have a new diagnosis of depression at the corresponding timepoint. | As the study progresses, the sample will increasingly consist of healthy adults that have never received a depression diagnosis. Psychological distress in the comparator group at each timepoint could reduce over time producing biased estimates, such as potentially inflated differences in psychological distress between adults with a recent diagnosis of depression and those without.<br>As the study progresses, previously reported depression diagnoses will be identified more effectively because there are a greater number of timepoints from which a history of can be observed. In comparison, there are fewer observations available to identify prior depression at earlier timepoints, and prior depression diagnoses might have been received prior to entering the UKHLS but not previously reported. The accuracy in identifying participants with prior depression therefore increases during the study, which could artificially lead to a possible decline in incidence over time. |

## Results

Across the nine timepoints, there were 344,337 observations of 66,623 adults. After taking into account whether there was available data regarding recent depression diagnoses, we analysed 341,764 observations of 66,360 adults. Of those, 93.4% had available data on the GHQ-12. At all timepoints, 55 to 56% of the sample were female and the mean age varied between 47 and 50 years (Table 4). At all timepoints, the minority of the sample were new entrants, however the proportion increased at timepoint 4 as a result of the Immigrant and Ethnicity Boost sample.

**Table 4.**
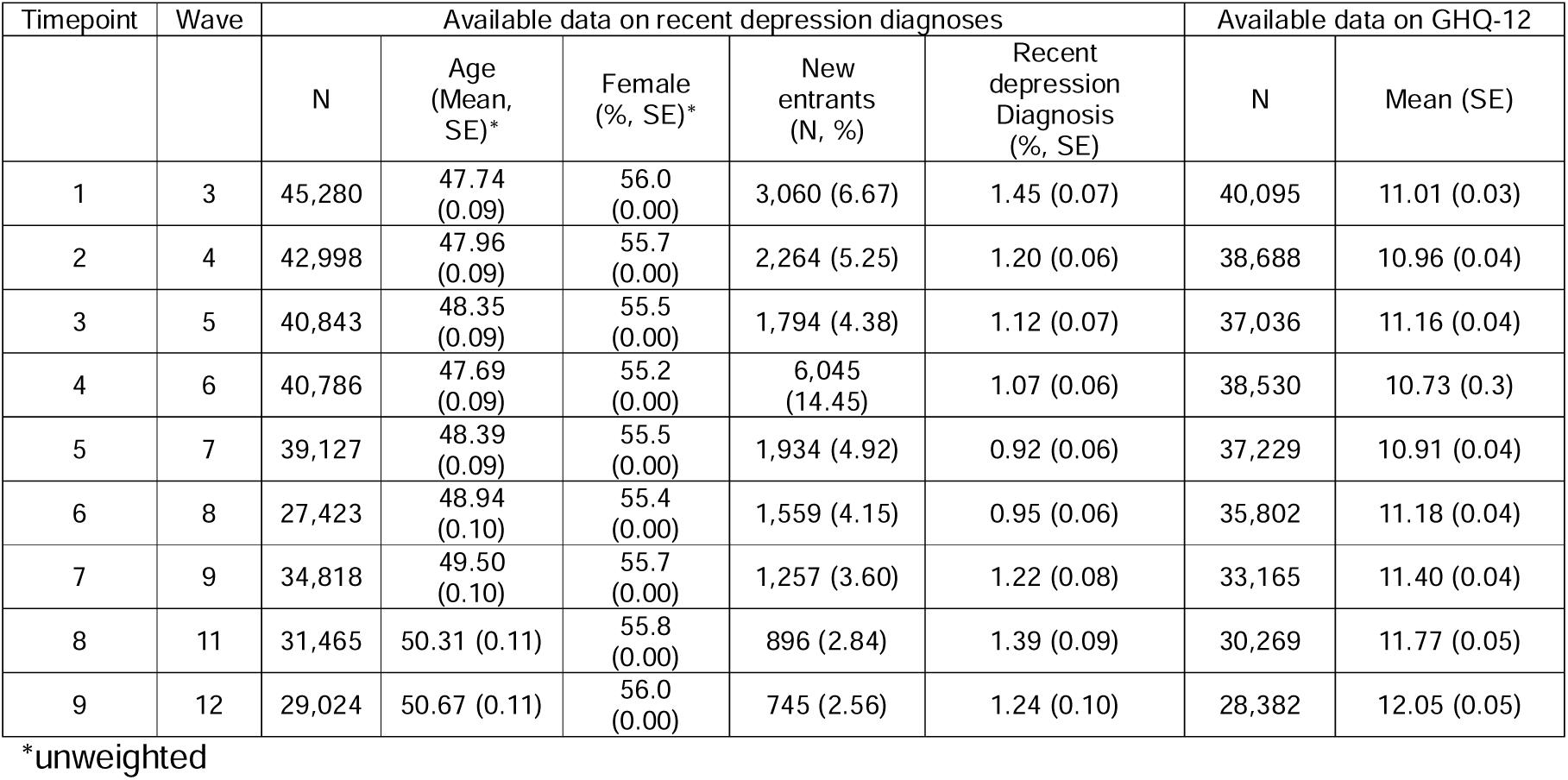
Adults with available data on recent depression diagnoses.

### Overall sample

From 2011-2013 to 2020-2022, between 3.53% (95% CI: 3.00% to 4.06%) and 4.96% (95% CI: 4.41% to 5.50%) of adults that met caseness for psychological distress reported a recent diagnosis of depression at that same timepoint (Figure 1a). Accordingly, approximately 95% of adults that reported high psychological distress did not recently receive a new diagnosis of clinical depression. In comparison, between 0.34% (95% CI: 0.26% to 0.42%) and 0.64% (95% CI: 0.54% to 0.74%) of adults that did not meet caseness for psychological distress reported a recent diagnosis of depression at that same timepoint. The proportion of the overall sample that reported a recent depression diagnosis was fairly stable over time (between 0.92% and 1.45%).

**Figure 1.**
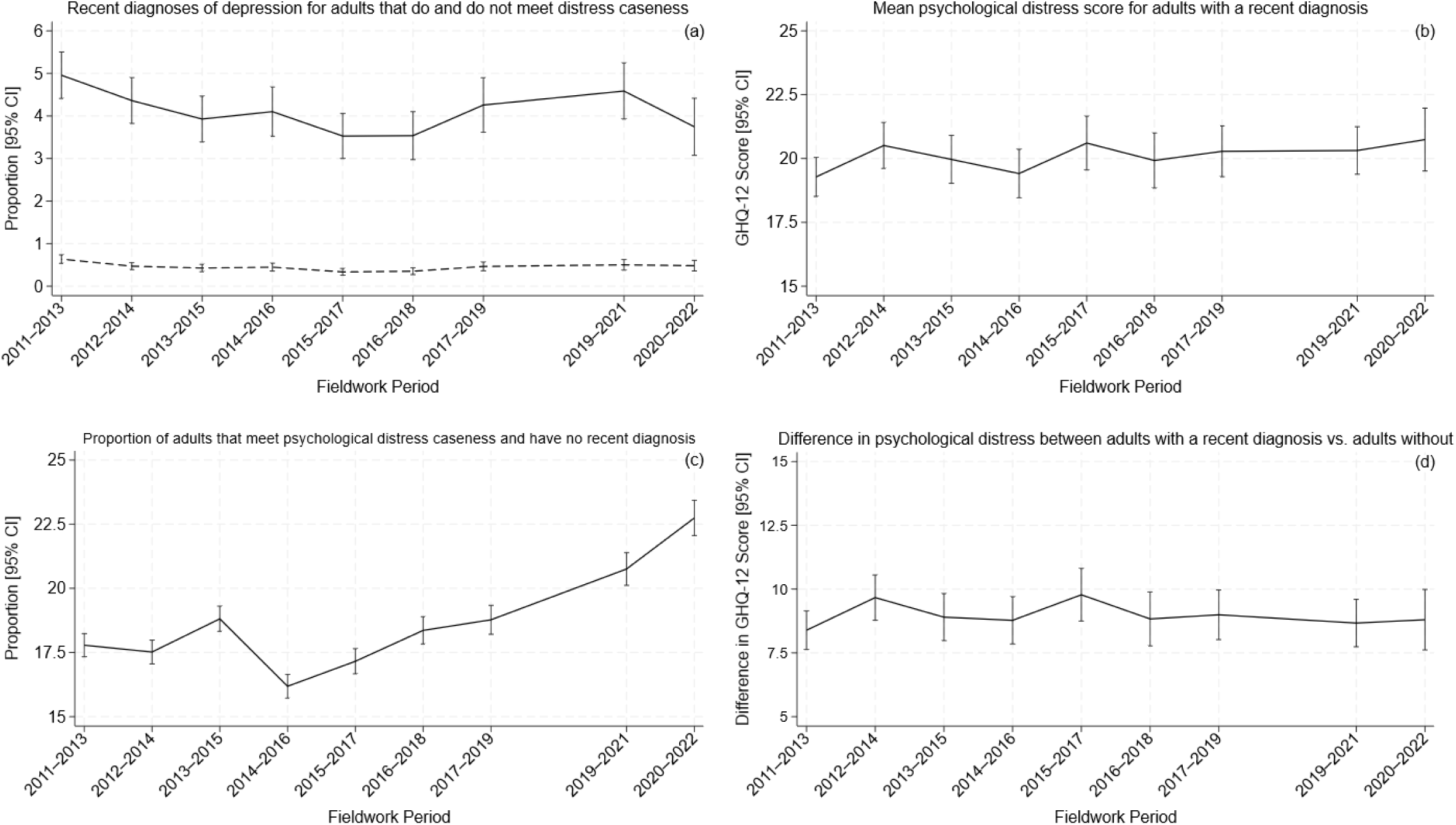
Time trends in depression diagnoses and psychological distress in the UK adult population. Note: In Figure 1a, the full line represents adults that report psychological distress according to the GHQ-12 and the dashed line represents adults that do not report psychological distress.

For adults with a recent diagnosis of depression, the severity of psychological distress (as indicated by mean GHQ-12 score) remained relatively stable from 2011-2013 to 2020-2022 (Figure 1b). The mean GHQ-12 score fluctuated between 19.27 at 2011-2013 (95% CI: 18.51 to 20.04) and 20.73 at 2020-2022 (95% CI: 19.51 to 21.97) and confidence intervals overlapped.

The proportion of the adult population in the UK that meet caseness for psychological distress but have not had a recent diagnosis of depression gradually rose from 17.78% in 2011-2013 (95% CI: 17.33% to 18.24%) to 22.74% in 2020-2022 (95% CI: 22.05% to 23.43%), with a small reduction in observed in 2014-16 (Figure 1c). Across time, adults with a recent diagnosis of depression had on average scores on the GHQ-12 between 8 and 10 points greater than adults without a recent diagnosis, with the difference stable from 2016-2018 (*b* = 8.83 [95% CI: 7.77 to 9.88]) to 2020-2022 (*b*= 8.80 [95% CI: 7.61 to 9.98]) (Figure 1d).

### Sex

Throughout all timepoints, recent depression diagnoses were more common in women than men (Figure 2a). The proportion of women that reported a recent depression diagnosis decreased from 2011-2013 (1.93% [95% CI: 1.73% to 2.13%]) to 2016-2018 (1.08% [95% CI: 0.91% to 1.25%]) but increased in the following years (2017-2022). Rates of new diagnosis of depression appear to be more stable over time for men, but there has been an increase in the proportion of men reporting a depression diagnosis from 2015-2017 (0.64% [95% CI: 0.49% to 0.78%]) to 2019-2021 (1.12% [95% CI: 0.88% to 1.36%]). For men and women with a recent diagnosis of depression, the severity of psychological distress reported has remained relatively stable from 2011-2013 to 2020-2022 (Figure 2b). Furthermore, there is no clear evidence that the severity of psychological distress in adults with a recent diagnosis of depression varied by sex.

**Figure 2.**
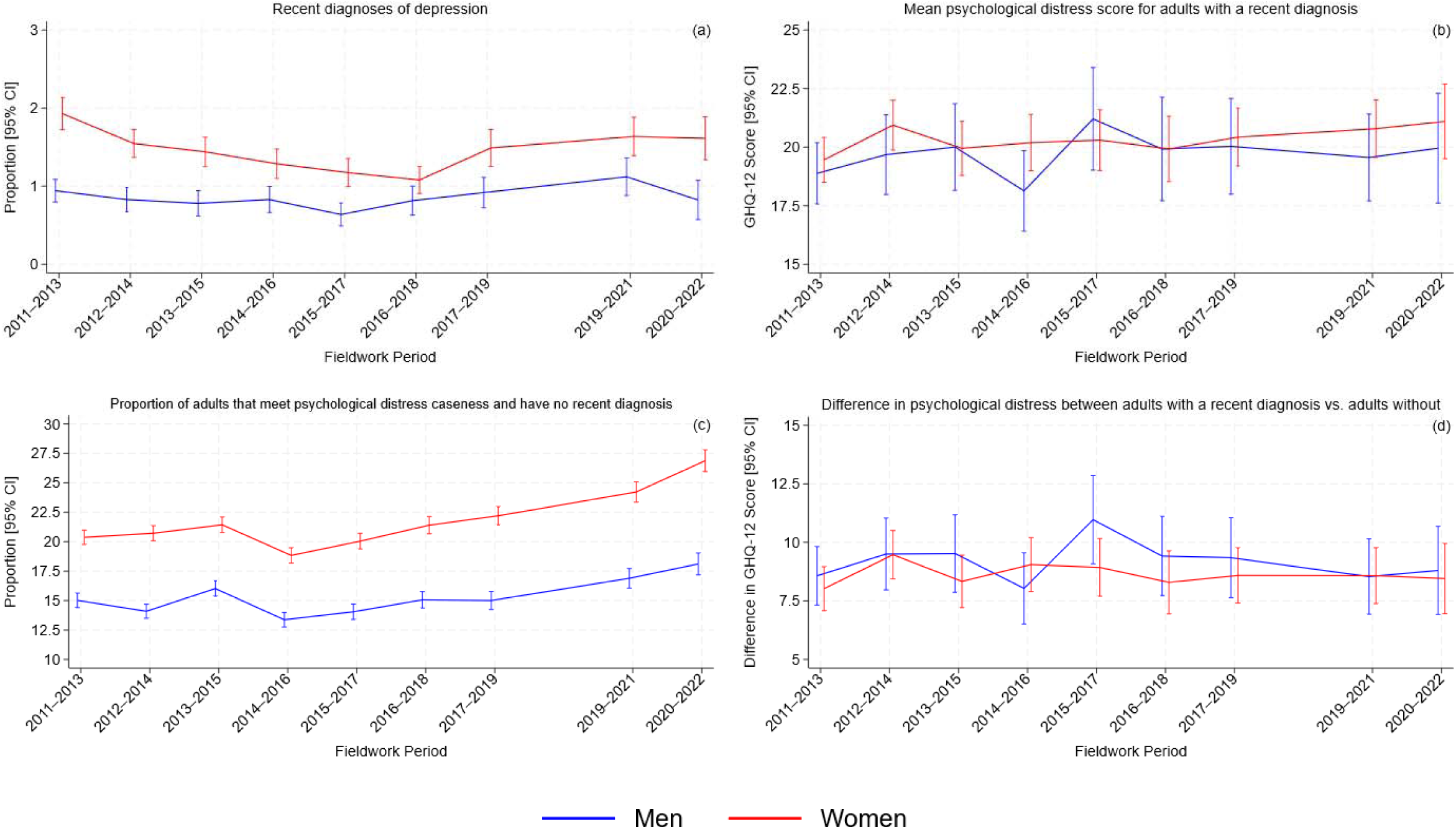
Time trends in depression diagnoses and psychological distress in the UK adult population, stratified by sex.

At all timepoints, women were more likely to meet caseness for psychological distress and not have a recent diagnosis of depression compared to men (Figure 2c). The proportion of men reporting psychological distress without a recent diagnosis slightly increased from 15.01% in 2011-2013 (95% CI: 14.40% to 15.62%) to 18.11% in 2020-2022 (95% CI: 17.18% to 19.04%). This increase has been larger for women (20.37% in 2011-2013 [95% CI: 19.76% to 20.98%], 26.87% in 2020-2002 [95% CI: 25.95% to 27.80%]). The difference in psychological distress between adults with and without a recent depression diagnosis is a similar size for both men and women at most timepoints, and this difference remained relatively stable over time for either sex (Figure 2d).

### Age

Of the four age groups, a clear increase in the proportion of adults reporting a recent depression diagnosis was only seen for those aged 16 to 24 (Figure 3a). This increase is apparent from 2016-2018 (1.45% [95% CI: 1.01% to 1.89%]) to 2020-2022 (2.02% [95% CI: 1.24% to 2.80%]). For all age groups, the severity of psychological distress in adults with recently diagnosed depression has remained relatively stable from 2011-2013 to 2020-2022 (Figure 3b). Potential differences in the severity of distress across age groups is obscured by large standard errors, particularly for the 16-24 group at later timepoints.

**Figure 3.**
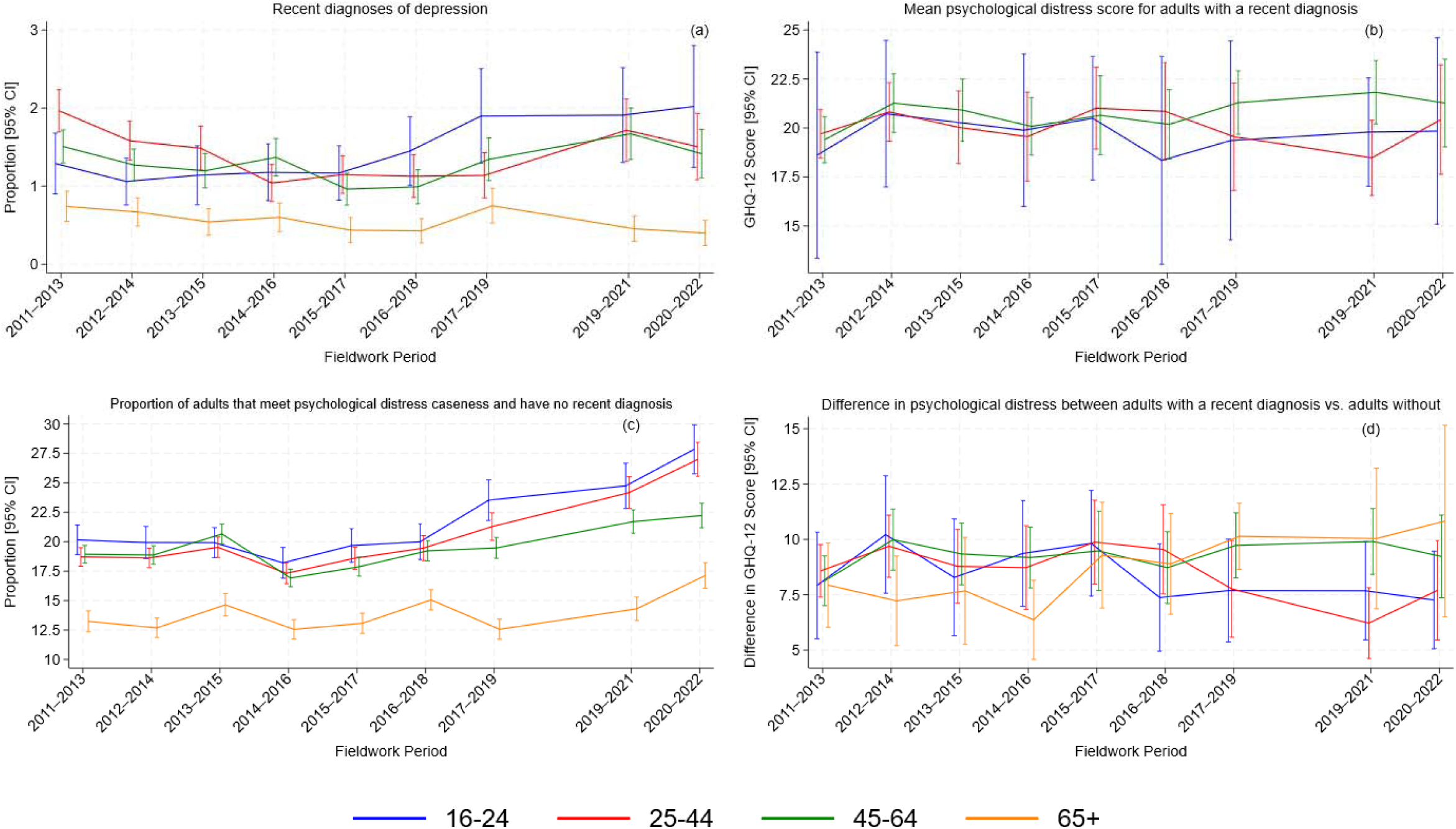
Time trends in depression diagnoses and psychological distress in the UK adult population, stratified by age. Note: Analyses of mean GHQ-12 score for adults aged 65+ recently diagnosed with depression are not included in Figure 3b as there were insufficient design degrees of freedom to estimate confidence for 4 of the 9 estimates, and the estimate at 2015-2017 (95% CI: 3.95 to 34.79) has large confidence intervals that reduce the readability of the figure.

At all timepoints, adults aged 65 or older were less likely to report psychological distress without a recent diagnosis to the other age groups (Figure 3c). For all age groups, the proportion of adults that report psychological distress but do not have a recent diagnosis of depression has risen over time. This increase is most apparent for adults between the ages of 16 and 44. This has coincided with a smaller difference in psychological distress between adults with and without a recent depression diagnosis for those aged 16 to 24 and 25 to 44 over recent years, whereas this difference in distress according to diagnosis appears consistent over the final timepoints for the older age groups (Figure 3d).

### Age and sex

When stratified by age and sex, changes in the proportion of the sample that report a recent depression diagnosis are most clear for women aged 16 to 24, where prevalence ranged between 1.6% to 1.9% from 2011-2013 to 2016-2018, before increasing to between 2.4% and 2.6% from 2017-2019 to 2020-2022 (Figure 4a). For men, there is very little age difference in the proportion reporting distress without a recent depression diagnosis, except for men aged 65 or older where the proportion is consistently lower and there is little change over time (Figure 4b). Distress without a diagnosis was more common in women aged 16 to 24. Across both sexes and all age groups, the proportion reporting psychological distress without a recent depression diagnosis appeared to have increased the most for women aged 16 to 44. Owing to large standard errors, the difference in psychological distress between men with and without a recent depression diagnosis is hard to compare between different age groups (Figure 4c). The difference in psychological distress between adults with and without a recent depression diagnosis has decreased the most for women aged 16-24, where it ranged from 7.8 to 9.2 in the first five timepoints, but between 6 and 6.6 in the final four timepoints.

**Figure 4.**
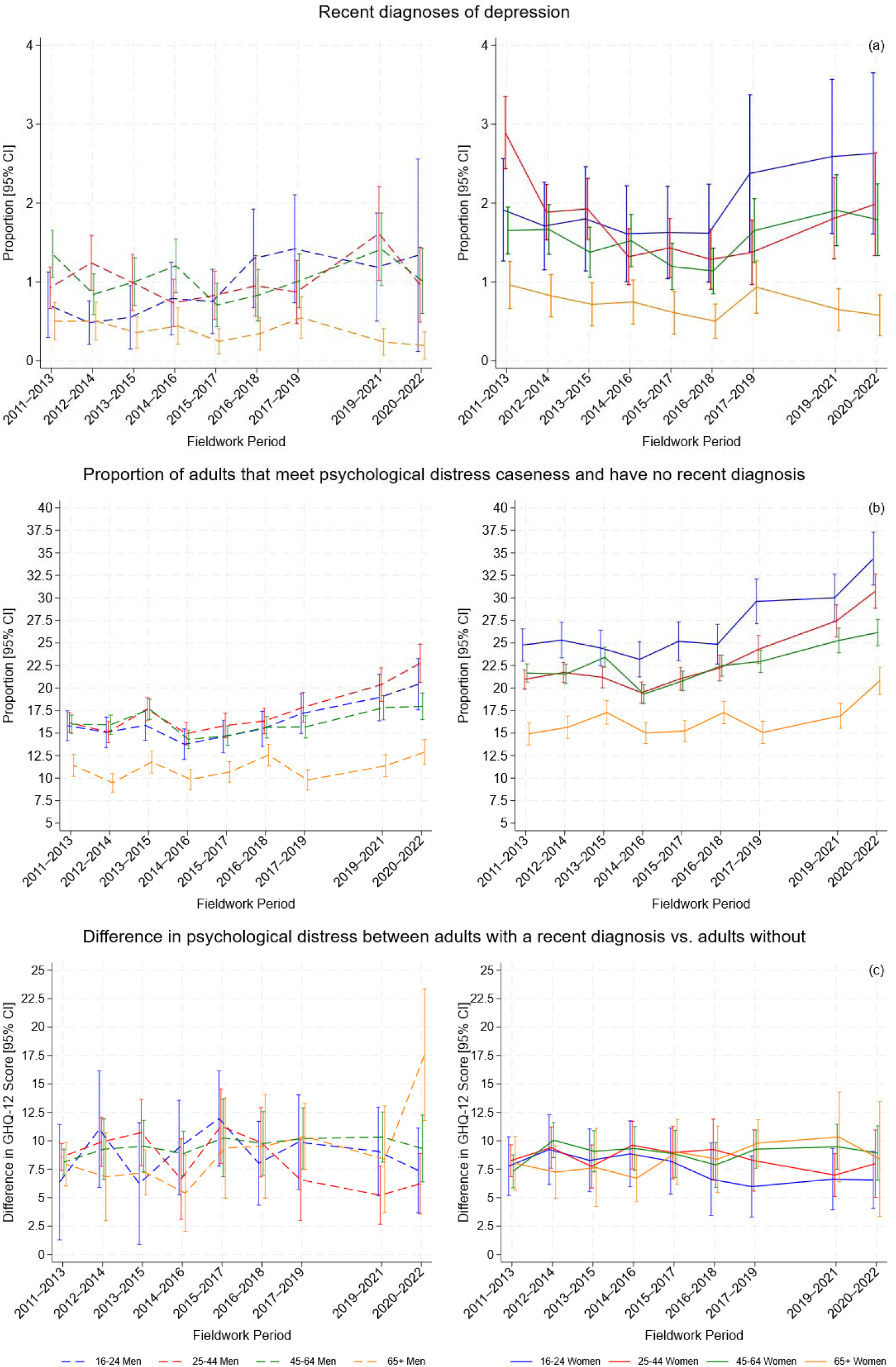
Time trends in depression diagnoses and psychological distress in the UK adult population, stratified by age and sex. Note: Analyses of mean GHQ-12 score for adults recently diagnosed with depression are not included as there were insufficient design degrees of freedom to estimate confidence for 26 of the 72 estimates.

### Cohort

There has been a decrease in the proportion of all cohorts reporting new diagnoses of incident depression from 2011-2013 to 2015-2017 or 2016-2018. From then, there is an apparent increase for Millennials and Generation X only. For Millennials, the proportion of the sample with newly diagnosed depression was 1.11% in 2015-2017 (95% CI: 0.83% to 1.39%), above 1.4% in all following timepoints and highest in 2019-2021 (1.87% [95% CI: 1.39% to 2.34%]). For Generation X, the proportion of the sample with newly diagnosed depression was 1.01% in 2016-2018 (95% CI: 0.76% to 1.27%), above 1.3% in all following timepoints and highest in 2019-2021 (1.79% [95% CI: 1.41% to 2.15%]). Across all cohorts, the severity of psychological distress in adults recently diagnosed with depression has remained relatively stable over time (Figure 3b). However, in recent years, findings suggests more severe psychological distress in Generation X (mean GHQ-12 in 2020-2022 = 22.0 [95% CI: 20.13 to 23.81]) and Baby Boomers (21.3 [95% CI: 18.85 to 23.65]) with a recent diagnosis of depression compared to Millennials (18.26 [95% CI: 15.75 to 20.76]) (Figure 5b).

**Figure 5.**
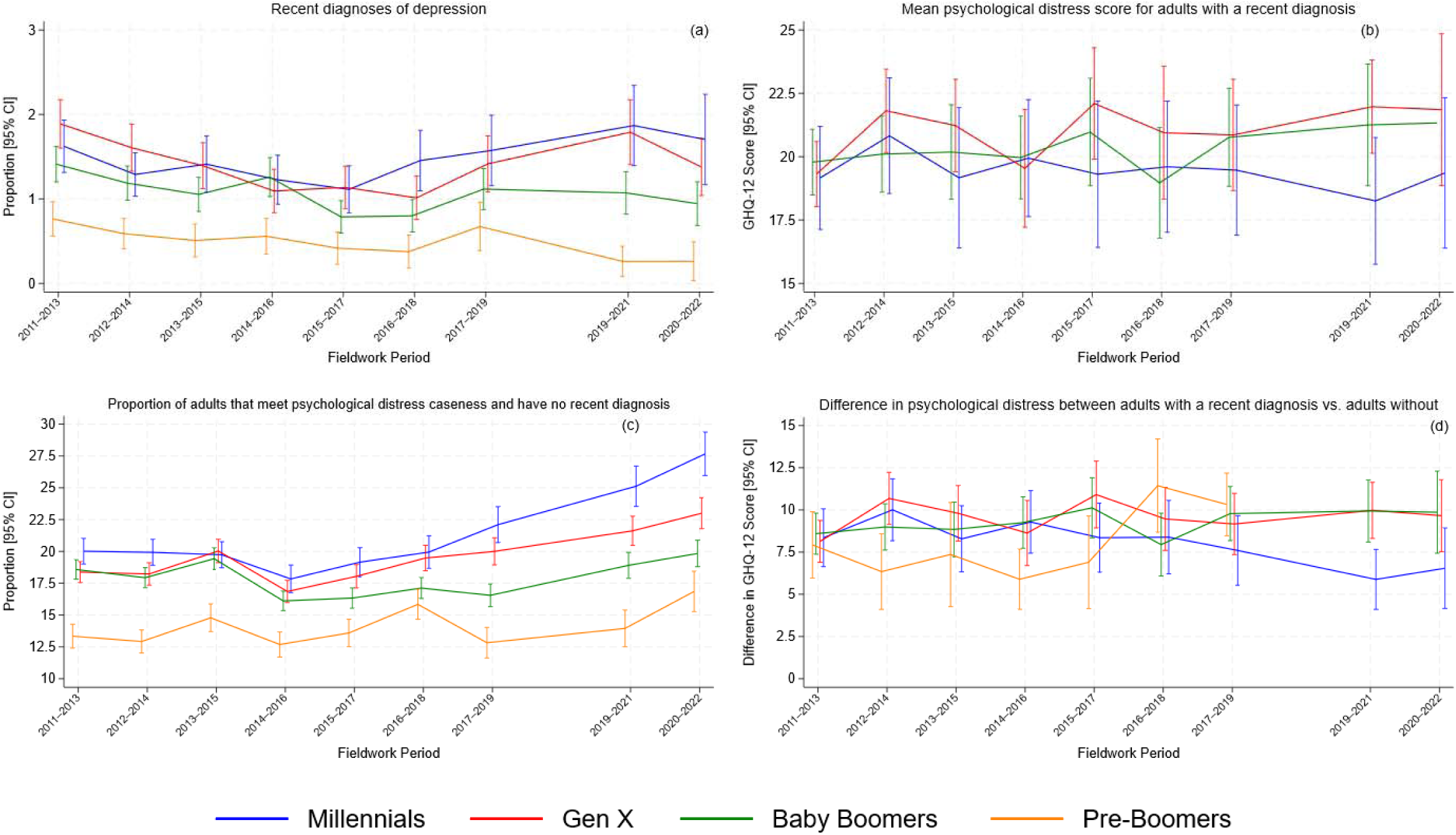
Time trends in depression diagnoses and psychological distress in the UK adult population, stratified by cohort. Note: Analyses of mean GHQ-12 score for Pre-Boomers recently diagnosed with depression are not included in 5b as there were insufficient design degrees of freedom to estimate confidence intervals for 7 of the 9 estimates. The estimates for Pre-Boomers at 2019-2021 (95% CI: -0.27 to 14.34) and 2020-2022 (95% CI: -1.28 to 21.66) are excluded from 5d due to large confidence intervals that reduce the readability of the figure.

From 2011-2013 to 2014-2016, the proportion of the sample reporting psychological distress without a recent depression diagnosis is similar between Baby Boomers, Generation X and Millennials (Figure 5c). From 2014-2016, an increase is apparent for Generation X and Millennials, whilst the rate remains relatively stable for Baby Boomers. Across all timepoints, Pre-Boomers are less likely to report experiencing distress without a recent diagnosis than other cohorts. The difference in psychological distress between adults with and without a recent depression diagnosis has remained relatively stable over time for Generation X and Baby Boomers (Figure 5d). The difference in psychological distress between Pre-Boomers with and without a recent depression appears smaller than other cohorts from 2012-2014 to 2015-2017, before increasing in the two following timepoints. The difference in psychological distress between Millennials with and without a recent depression diagnosis has decreased over the study period, reducing from 8.34 (95% CI: 6.64 to 10.06) at the first timepoint, 2011-2013, to 6.54 (95% CI: 4.16 to 8.93) at the final timepoint, 2020-2022.

### Ethnicity

Throughout all timepoints, White adults more commonly reported a new diagnosis of depression compared to ethnic minority adults (Figure 6a). In contrast, ethnic minority adults were more likely to meet caseness for psychological distress and not have a recent diagnosis of depression compared to White adults, most clearly from 2011-2013 to 2014-2016 (Figure 6c). Since 2015-2017, increases in the proportion reporting psychological distress without a recent depression diagnosis has been comparable over time for both White and ethnic minority adults.

**Figure 6.**
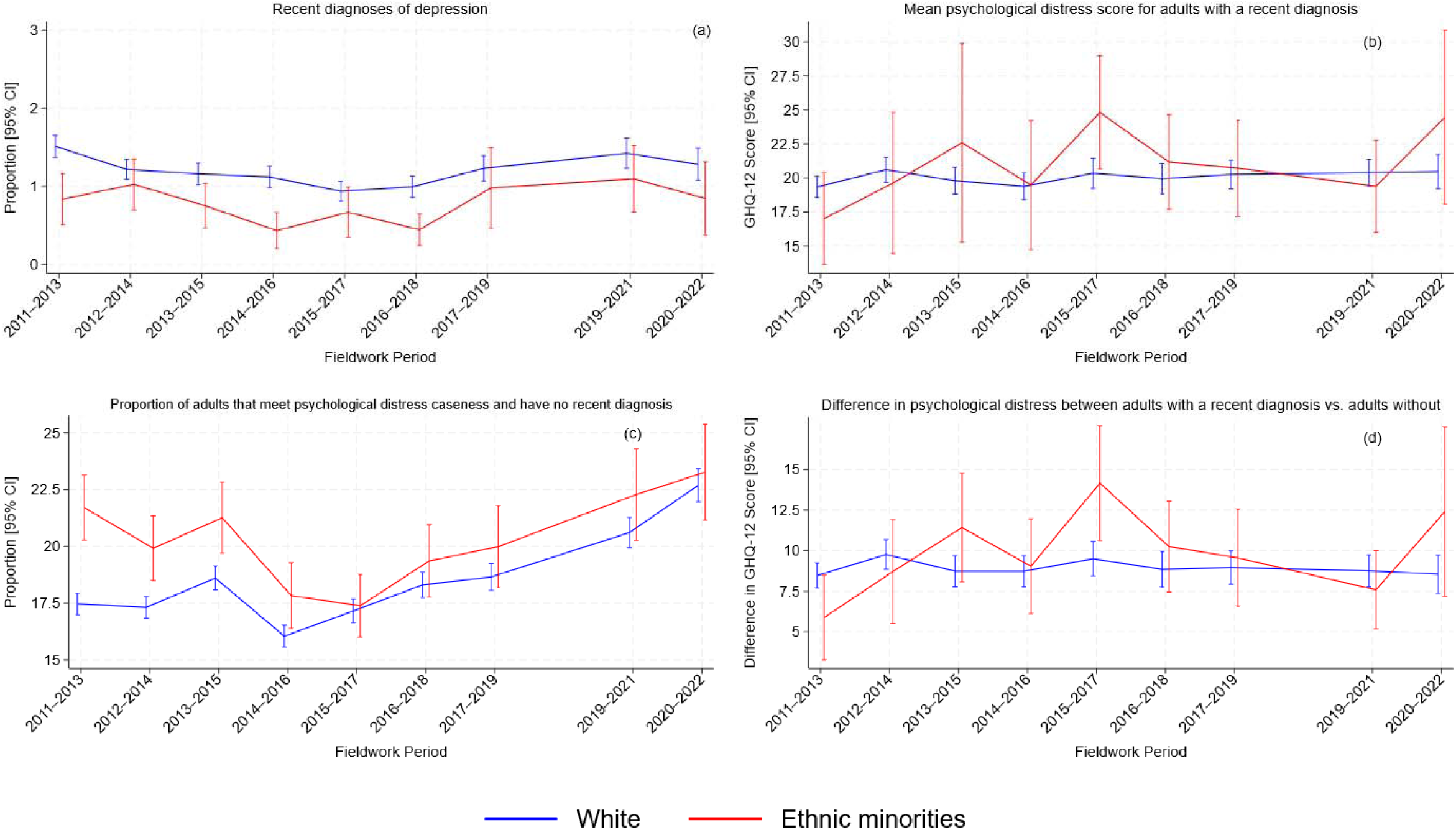
Time trends in depression diagnoses and psychological distress in the UK adult population, stratified by ethnicity (binary classification).

For White adults with a recent diagnosis of depression, the severity of psychological distress has remained relatively stable from 2011-2013 to 2020-2022 (Figure 6b). The severity of psychological distress for ethnic minority adults with a recent diagnosis of depression appears to have varied over time, however large standard errors prohibit strong conclusions regarding long-term trends. The difference in psychological distress between White adults with and without a recent depression diagnosis has remained steady over time, whilst this has fluctuated over time for ethnic minority adults, but changes are at likely to be at least partly random fluctuations driven by the high level of uncertainty at many timepoints (Figure 6d).

In analyses where ethnicity was classified using four groups (White, Mixed, Asian and Black), trends in diagnosed depression were relatively constant except for adults of mixed ethnicity (Figure 7). At most timepoints, a greater proportion of mixed ethnicity adults reported psychological distress without a recent diagnosis of depression compared to other ethnicities. The difference in psychological distress between Asian, Mixed and Black adults with and without a recent depression diagnosis fluctuated over time, however changes may be spurious fluctuations driven by the high level of uncertainty of estimates (Figure 7d).

**Figure 7.**
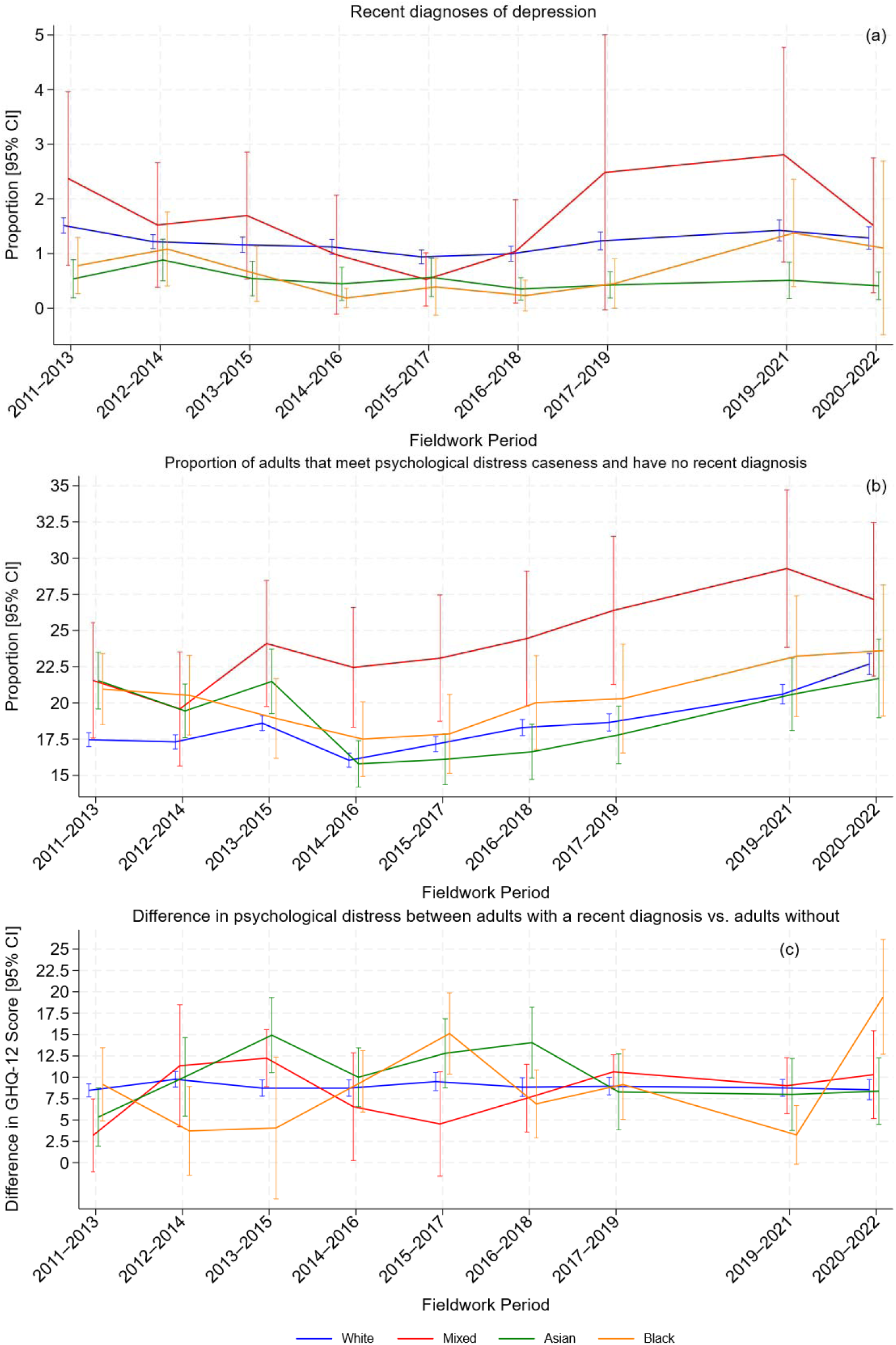
Time trends in depression diagnoses and psychological distress in the UK adult population, stratified by ethnicity (four group classification). Note: Analyses of mean GHQ-12 score for adults recently diagnosed with depression are not included as there were insufficient design degrees of freedom to estimate confidence intervals for 13 of the 36 estimates.

### Education level

There was a general trend where a smaller proportion of adults with higher education qualifications reported incident depression compared to adults that do not have higher education qualifications from 2011-2013 to 2015-2017 (Figure 8a). Since then, recent diagnoses of depression in adults with upper or lower secondary qualifications have increased, whilst there has been a decline for adults with no qualifications (1.63% [95% CI: 1.26% to 2.01%] in 2011-2013, 0.67% [95% CI: 0.24% to 1.09%] in 2020-2022). Trends in the level of psychological distress for adults with a recent diagnosis of depression were relatively consistent for all education levels (Figure 8b).

**Figure 8.**
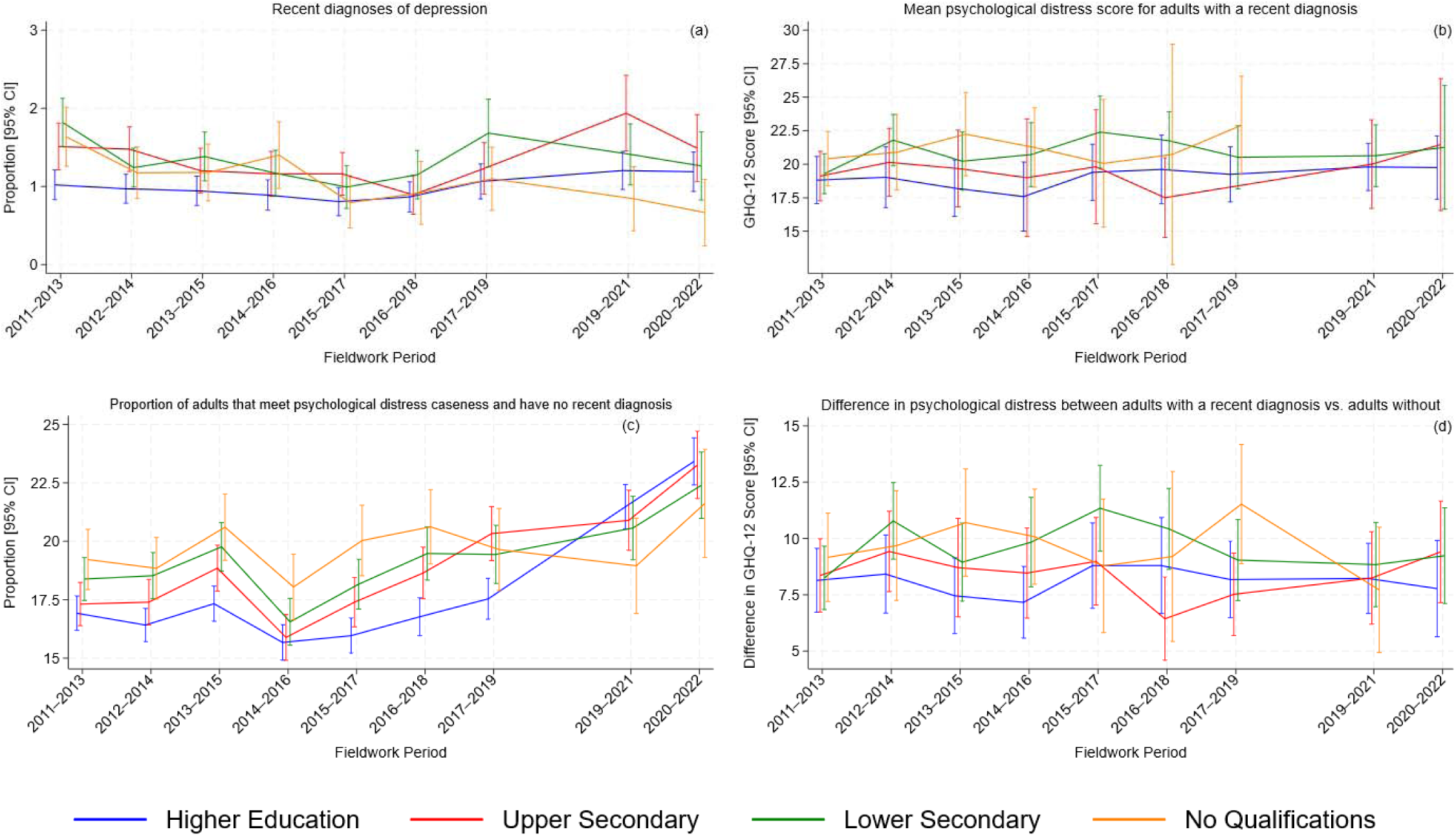
Time trends in depression diagnoses and psychological distress in the UK adult population, stratified by highest educational qualification attained. Note: The estimates for the no qualifications group at timepoint 2019-2021 (95% CI: 1.37 to 37.10) and timepoint 2020-2022 are excluded from 8b due to large confidence intervals, and insufficient design degrees of freedom to estimate confidence intervals respectively. The estimate for the upper secondary qualifications group at waves timepoint 2017-2019 (95% CI: 7.19 to 30.88) is excluded from 8b due to large confidence intervals. The estimate for the no qualifications group at timepoint 2020-2022 (95% CI: 3.58 to 18.86) are excluded from 8d due to large confidence intervals.

Cases of experiencing psychological distress without receiving a recent diagnosis of depression were initially lowest for adults with higher education qualifications (16.92% [95% CI: 16.18% to 17.66%] in 2011-2013), but by 2020-2022 cases were highest in adults with higher education qualifications (23.42% [95% CI: 22.41% to 24.42%]) (Figure 8c). In comparison, the shallowest increase was observed for adults with no qualifications (19.21% [95% CI: 17.93% to 20.51%] in 2011-2013, 21.62% [95% CI: 19.31% to 23.93%] in 2020-2022).

The difference in psychological distress between adults with and without a recent depression diagnosis was smallest for adults with higher education qualifications in the early timepoints (Figure 8d). There are clear changes over time across all educational groups, but these changes might be random fluctuations owing to the high level of uncertainty of most estimates, and there are no overly clear patterns for any group.

### Financial stress

At all timepoints, adults experiencing high or moderate levels of financial stress were more likely to have a recent diagnosis of depression than adults with no financial stress (Figure 9a). Whilst rates of new depression diagnoses have remained mostly stable for adults with no or moderate financial stress, the proportion of the sample reporting incident depression has increased for adults with high levels of financial stress from 2.91% in 2016-2018 (95% CI: 2.07% to 3.75%) to 3.99% in 2020-2022 (95% CI: 2.33% to 5.65%). At most timepoints, adults with recently diagnosed depression reported more severe psychological distress if they were experiencing higher levels of financial stress, with time trends relatively consistent for each of the three subgroups (Figure 9b).

**Figure 9.**
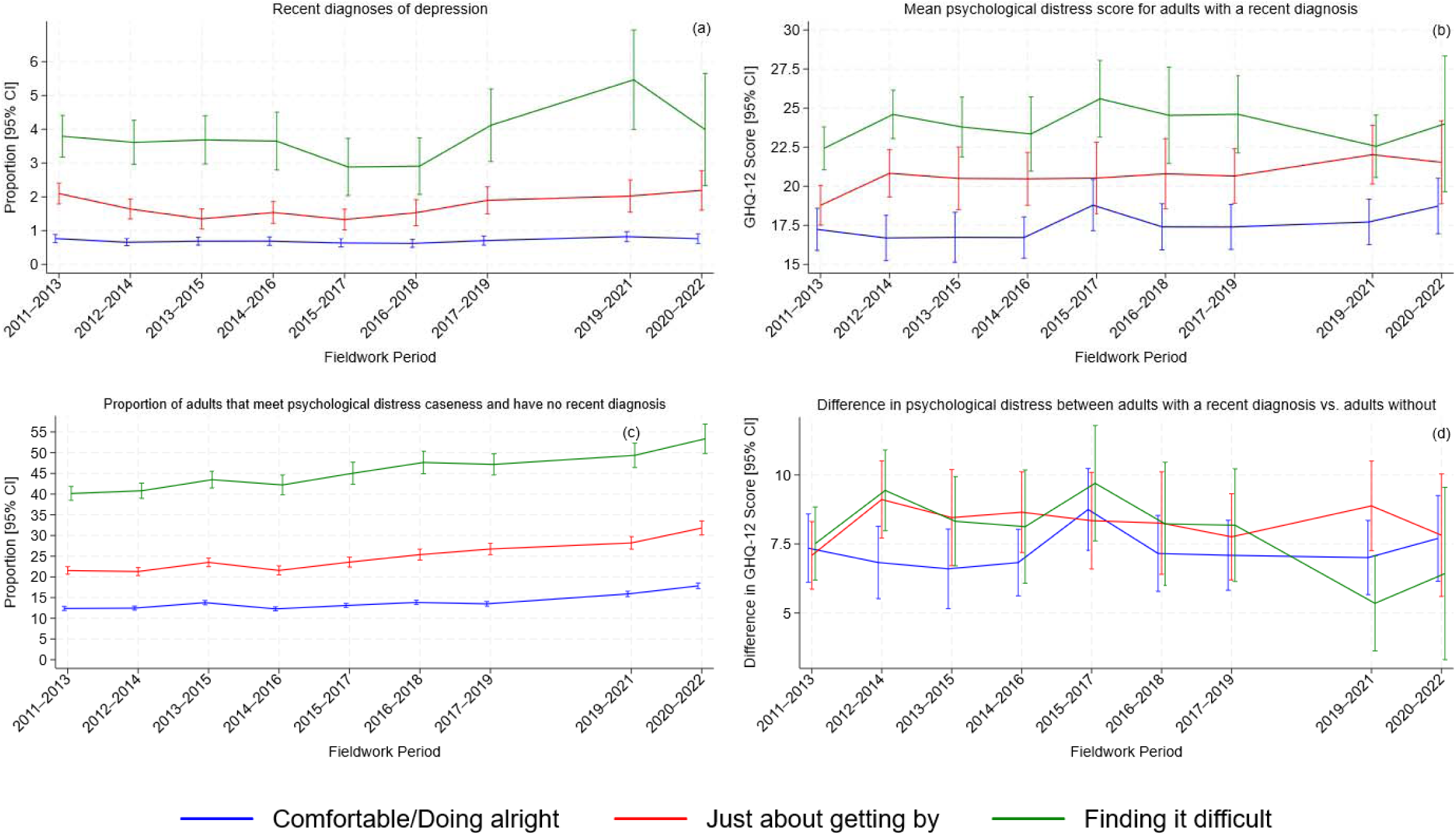
Time trends in depression diagnoses and psychological distress in the UK adult population, stratified by financial stress.

The proportion of the sample reporting psychological distress without a recent diagnosis of depression was high at all timepoints for adults with high levels of financial stress, with an increase seen from 2011-2013 (40.15% [95% CI: 38.49% to 41.82%]) to 2020-2022 (53.35% [95% CI: 49.81% to 56.90%]) (Figure 9c). The proportion of the sample reporting psychological distress without a recent diagnosis of depression has also risen over time for adults experiencing moderate or no financial stress, but at a slower rate. The difference in psychological distress between adults with and without a recent depression diagnosis has remained steady over time regardless of the level of financial stress experienced (Figure 9d). At earlier timepoints the difference appears smaller for adults with no financial stress compared to adults experience high or moderate financial stress. For adults experiencing high levels of financial stress there is a sharp decline from 2017-2019 (*b*=8.18 [95% CI: 6.14 to 10.22]) to 2019-2021 (*b*=5.35 [95% CI: 3.64 to 7.07]), before rebounding slightly in 2020-2022 (*b*=6.43 [95% CI: 3.31 to 9.55]).

### Excluding adults who reported previous depression diagnoses

After excluding adults who reported previous depression diagnoses, the proportion of the sample that reported a new diagnoses of depression decreased compared to the main analysis, suggesting that some adults reported a new diagnosis of depression multiple times at different timepoints (Figure 10a). In these analyses, between 2.06% (95% CI: 1.53% to 2.60%) and 4.15% (95% CI: 3.61% to 4.69%) of adults that met caseness for psychological distress reported a recent diagnosis of depression at that same timepoint, compared to between 3.53% and 4.96% in the main analyses. The proportion reporting new diagnoses of depression remained fairly stable, consistent with the main analysis, until the final timepoint were there was a clear drop.

**Figure 10.**
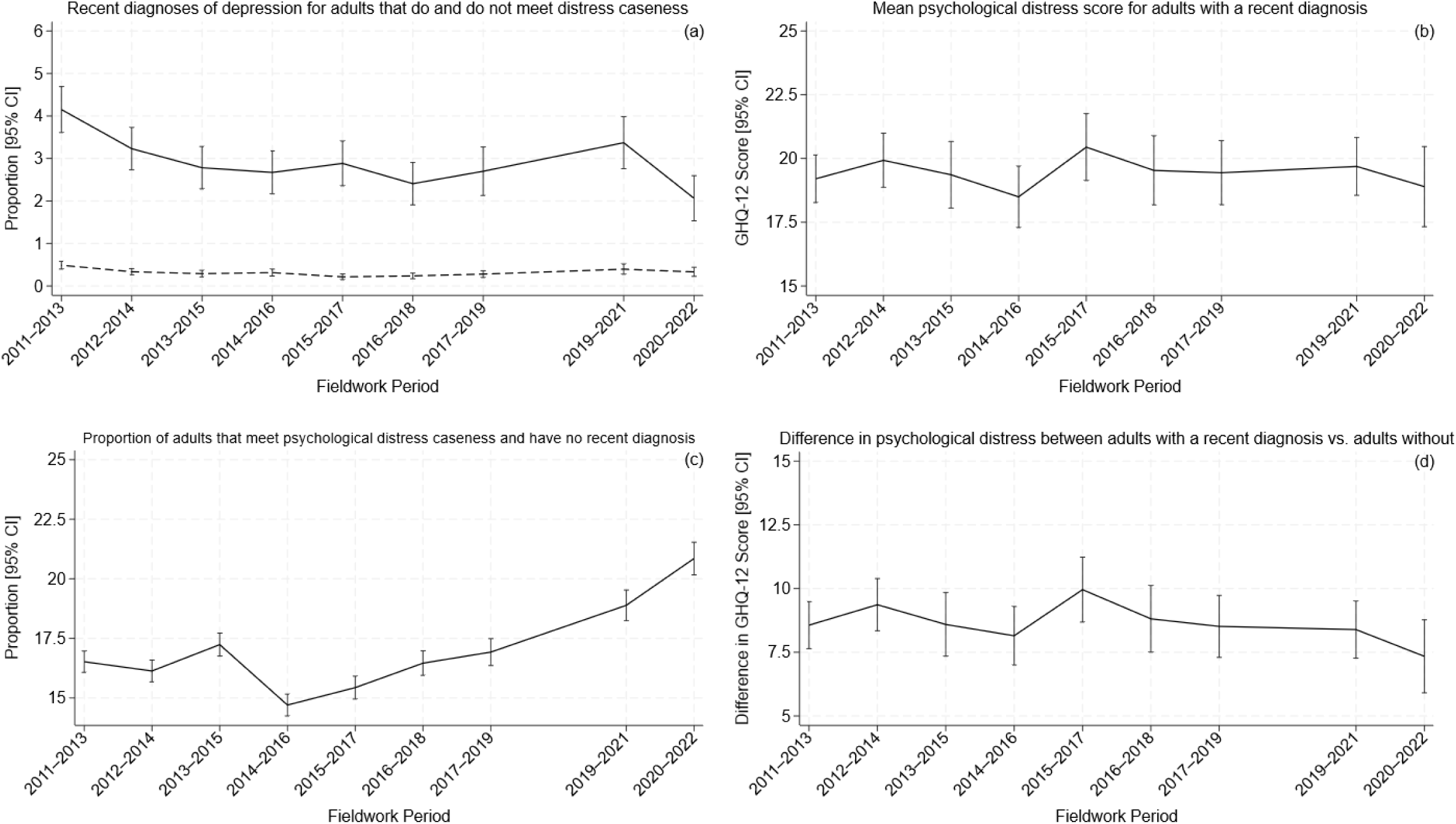
Time trends in depression diagnoses and psychological distress in the UK adult population, excluding adults from future timepoints after first reporting a diagnosis of depression.

There was still a clear trend of an increasing proportion of the adult population that meet caseness for psychological distress but do not have a recent diagnosis of depression (16.52% [95% CI: 16.07% to 16.97%] in 2011-2013 to 20.85% [95% CI: 20.17% to 21.53%] in 2020-2022) (Figure 10c). At all timepoints, the proportion reporting psychological distress without a recent depression diagnosis was approximately two percentage points smaller than in the main analyses, however the general trend over time remained the same.

The level of psychological distress reported by adults with a recent diagnosis of depression (Figure 10b) and the difference in psychological distress between adults with a recent diagnosis and adults without has (Figure 10d) remained stable across the study period.

The trends identified from analyses stratified by different sociodemographic characteristics were in most cases consistent with one another (Supplementary Figures 1 to 8). Across timepoints for all subgroups, the proportion reporting a recent diagnosis of depression was less compared to the main analyses. In analyses stratified by age group, decreases in the proportion of the sample reporting a recent diagnosis of depression were less evident for 16- to 24-year-old adults, likely due to a short period of risk for previous depressive episodes, and therefore less likely of reporting a previous diagnosis of depression at an earlier wave. Furthermore, the proportion of 25- to 65-year-olds reporting psychological distress without a recent depression diagnosis was less compared to the main analyses, whilst there was minimal change for adults aged 16 to 24 (Supplementary Figure 2). As such, an increase over time in the proportion reporting psychological distress without a recent depression diagnosis was only clear seen for adults aged 16 to 44, and there is less evidence of an increase for adults aged 45 to 64.

## Discussion

In this large study of the general adult population in the UK, we described time trends in recently diagnosed depression and its relationship with psychological distress. We found that the incidence of diagnosed depression has not increased over time, whilst cases of psychological distress without a diagnosis of depression have. The level of psychological distress experienced by adults recently diagnosed with depression has slightly increased over time, whilst the difference in psychological distress experienced by adults with and without a recent depression diagnosis had remained stable.

From 2009-2011 to 2020-2022, the incidence of depression diagnoses in the UK was stable across time, contrasting to an increase in common mental disorder recorded in primary care over a similar study period.^12^ We did observe changes in the incidence of depression in subgroups, which was most apparent for women between the ages of 16 and 24, replicating trends observed across different data sources in the UK.^1,12,13^ In our sample, the proportion of adults with a recent depression diagnosis and met psychological caseness was about five times greater than the proportion of adults with a recent depression diagnosis and that did not meet psychological caseness, indicating that reporting high levels of psychological distress corresponds to receiving a diagnosis over the study period.

The level of psychological distress experienced by adults newly diagnosed with depression remained consistent for the general population and for most subgroups. Consequently, this suggests that there has been little or no increase over time in the likelihood that adults with mild, transient or sub-threshold depression receive a diagnosis, as if this were the case we would expect the level of psychological distress to decline over time. On the contrary, the severity of psychological distress for adults recently diagnosed with depression slightly increased over time. Our study suggests that changes in the incidence of depression over time are unlikely to be primarily due to changes in help-seeking behaviours or diagnostic practices, such as lower thresholds where diagnoses are given.

We found that a substantial proportion of adults experience psychological distress without being recently diagnosed with depression, with this increasing over time. The rate of this increase in psychological distress has not been matched by increases in the incidence of diagnosed depression. It is important to note that for many of the adults in our study reporting psychological distress, (a) cases might be mild, transient or not warrant intervention,^14^ (b) they might have a recent diagnosis of another mental health problem, or (c) in the main analyses, had a past diagnosis of depression. However, the increasing trend over time suggests a rising mental healthcare gap in the UK. The proportion of adults with psychological distress but without a recent diagnosis, was highest in the final two timepoints. These timepoints overlap with the COVID-19 pandemic, which had a detrimental impact on population mental health and led to disruption in healthcare services which might have also delayed the receipt of diagnoses or treatment for mental health problems.^15^

Increases in the number of cases of psychological distress without a recent depression diagnosis were most apparent for women and younger cohorts, echoing previous analyses of five UKHLS waves.^16^ Ethnic minorities were less likely to report a recent diagnosis of depression compared to White adults, but ethnic minorities were more likely to experience psychological distress without a recent diagnosis compared to White adults across several timepoints. Accordingly, our findings suggest that ethnic minorities with psychological distress experience more barriers that prevent receiving a diagnosis of depression, adding to a large body of findings of ethnic inequalities in mental healthcare access in the UK.^7,17,18^ The difference in the proportion that report psychological distress without a recent diagnosis between White adults and ethnic minorities appears to have reduced over time, potentially indicating a reduction in the mental health care gap.

The difference in the level of psychological distress experienced between adults with a recent diagnosis of depression and adults without a recent diagnosis was stable over time in the general population, and for most subgroups. For most of the population, there is little evidence to suggest that any increase in the incidence of diagnosed depression is driven by changes in the likelihood that a diagnosis is given for a certain amount of distress. However, one of the largest changes observed over time was for women aged 16-24, where a decrease in the difference of psychological distress between adults with and without a recent depression diagnosis coincided with an increase in the incidence of depression diagnoses.

Overall, the trends identified, in addition to findings from other studies,^1–4^ might reflect worsening mental health in the population. It is has been argued that increases in self-reported distress could be due to changes in how people self-report over time, potentially driven by increased public awareness and reduced societal stigma of common mental health problems in UK society.^19,20^ The study period aligns with an increase in public programmes against mental health stigma, most notably, Time to Change that ran from 2007 to March 2021, that might have contributed to changes observed in public attitudes.^21^ However, our findings provide little support for the hypothesis that higher incidence of diagnosed depression does not indicate a decline in population mental health and instead might be due to increased healthcare seeking. We found little evidence that diagnoses were being received for less mild cases compared to earlier timepoints, meaning that any changes in healthcare seeking is unlikely to explain our findings. Overall, our study suggests that has been minimal changes in the incidence of depression in UK adults, and that there is also little evidence that changes in the incidence of depression diagnoses could be primarily driven by increases in healthcare seeking by those with less severe symptoms or a lowering of thresholds where diagnoses are given.

### Limitations

Our study has several limitations. We relied on self-reported diagnosis of depression, rather than health care records that might be more accurate (but are unavailable). However, our estimated incidence for new diagnoses of depression ranged from 0.92% to 1.45%, closely matching the incidence (1.4%) reported in 2024-2025 by NHS Digital, suggesting the measure used reflects the incidence observed in general practice.^22^ However, we cannot rule out measurement error and adults may have been inaccurately classed as receiving or not receiving a recent depression diagnosis.

We modelled new cases of depression based on reports of receiving a diagnosis in the past year or since the previous wave an adult attended, whilst psychological distress was measured later – specifically the date of completing the adult survey in the UKHLS. Accordingly, psychological distress reported at the survey timepoint might differ to the psychological distress experienced when the diagnosis was given. After receiving a diagnosis of, or treatment for depression, symptoms often improve in mild and moderate cases.^23^ As a result, our estimates of the level of psychological distress experienced by adults recently diagnosed might underestimate the true psychological distress experienced at the onset of depression and whilst a diagnosis is being sought. Additionally, an adult diagnosed with depression at an earlier timepoint but still experiences high psychological distress at later timepoints were considered as “distressed with no recent diagnosis” in timepoints that follow their diagnosis in the main analyses. Accordingly, the prevalence of adults that meet caseness but have not received a recent diagnosis of depression is likely to be an overestimate of the treatment gap in the UK adult population. However, the results from our main analyses were similar to results after excluding adults who had reported a depression diagnosis in a previous timepoint, suggesting that this did not substantively affect our findings. Given that the primary focus of the paper is change over time, these limitations should have little impact on the observed trends.

We estimated that almost one in four adults in the UK in 2020-2022 might be experiencing symptoms indicative of a mental health problem but do not have a recent diagnosis. This figure contrasts to that of the 2023-2024 Adult Psychiatric Morbidity Survey, which reported that only 2.8% of the adult population had an unmet request for mental health treatment in the past 12 months.^1^ Our measure, using the GHQ-12 caseness approach, is likely to capture distress, which in a number of cases might not be accompanied by sufficient impairment that would lead to a diagnosis or require treatment. Consequently, our estimates are unlikely to be a precise reflection of the level of unmet mental health needs in the adult population. However, the focus of our study is how any relationship has changed over time, and as this measure has increased over time, our study indicates that there might be an increase in the proportion of adults that experience high levels of psychological distress but don’t need, have not sought or are yet to receive treatment.

The size of the overall sample and the subgroups within it decreased over time due to study attrition. As a result, the precision of our estimates also decreased over time, potentially obscuring trends at the most recent timepoints. Our stratified analyses were also informed by sample size considerations, which might have led to the use of less informative categorisations of different social factors (e.g. ethnicity modelled as a binary variable rather than finer-grained groups), potentially hiding important time trends for specific groups. Concerns regarding power are most prominent for analyses that model recent diagnoses of depression, given the rarity of this outcome (less than 1.5% in the general population) and therefore the low number of cases within smaller subgroups. Finally, due to the design of the UKHLS our study timepoints overlap slightly with one another, not permitting finer grained year on year insights, however this has not impact on the broader time trends that are the focus of this study.

### Conclusion

The incidence of self-reported depression diagnoses in the UK has remained fairly stable over a decade, contrasting with an increase in the proportion of adults experiencing psychological distress yet do not report receiving a recent diagnosis of depression. Minimal changes over time in the level of distress experienced by adults recently diagnosed with depression, and the difference in psychological distress experienced by adults with and without a recent depression diagnosis, suggest that any changes in the incidence of depression diagnoses are unlikely to be primarily driven by increases in healthcare seeking or changes in diagnostician practices

## Data Availability

Data used in this study are available from the UK Data Service.

## Supplementary material

**Supplementary Figure 1.**
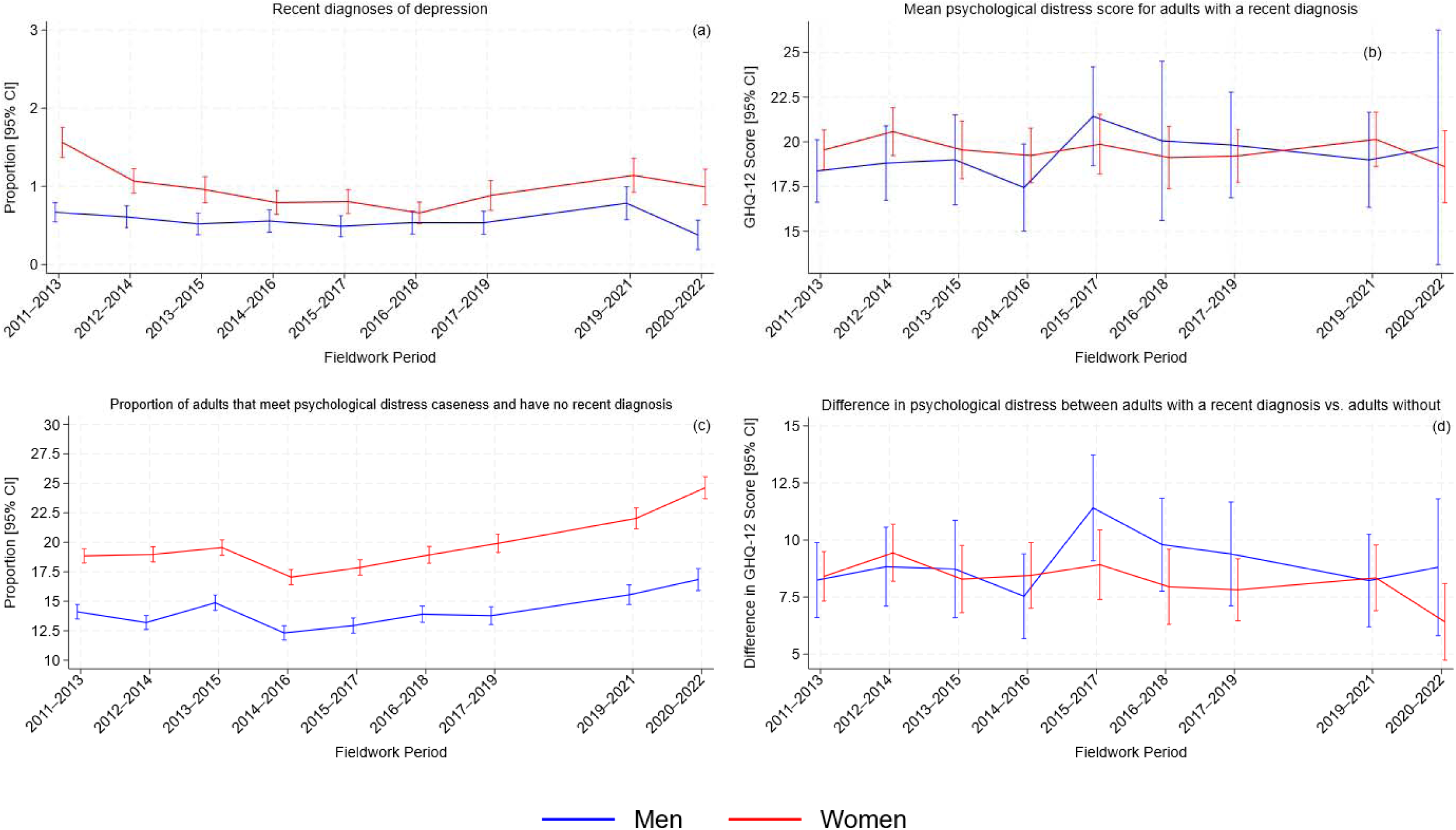
Time trends in depression diagnoses and psychological distress in the UK adult population stratified by sex, excluding adults from future timepoints after first reporting a diagnosis of depression.

**Supplementary Figure 2.**
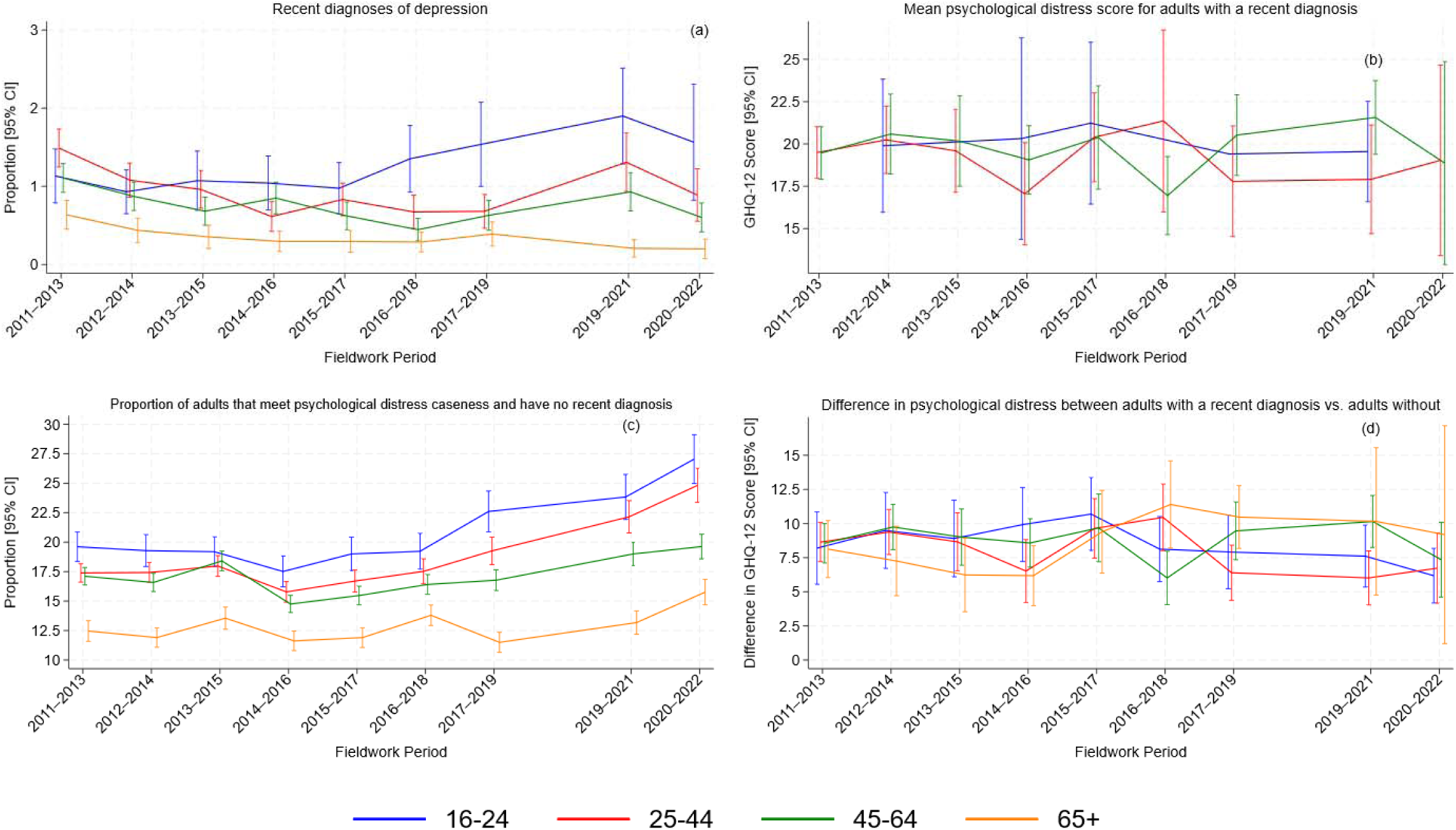
Time trends in depression diagnoses and psychological distress in the UK adult population stratified by age, excluding adults from future timepoints after first reporting a diagnosis of depression. Note: Analyses of mean GHQ-12 score for adults recently diagnosed with depression are not included in Supplementary Figure 2b for adults aged 65+ as there were insufficient design degrees of freedom to estimate confidence for 6 of the 9 estimates, and the estimate at 2017-2019 (95% CI: 5.63 to 35.21) has large confidence intervals that reduce the readability of the figure. The estimates for adults aged 16-24 at timepoints 2015-2017 and 2017-19, and at timepoints 2011-2013 (95% CI: 1.69 to 35.91) and 2020-2022 (95% CI: 5.92 to 31.24) are excluded from Supplementary Figure 2b due to insufficient design degrees of freedom to estimate confidence intervals and large confidence intervals respectively.

**Supplementary Figure 3.**
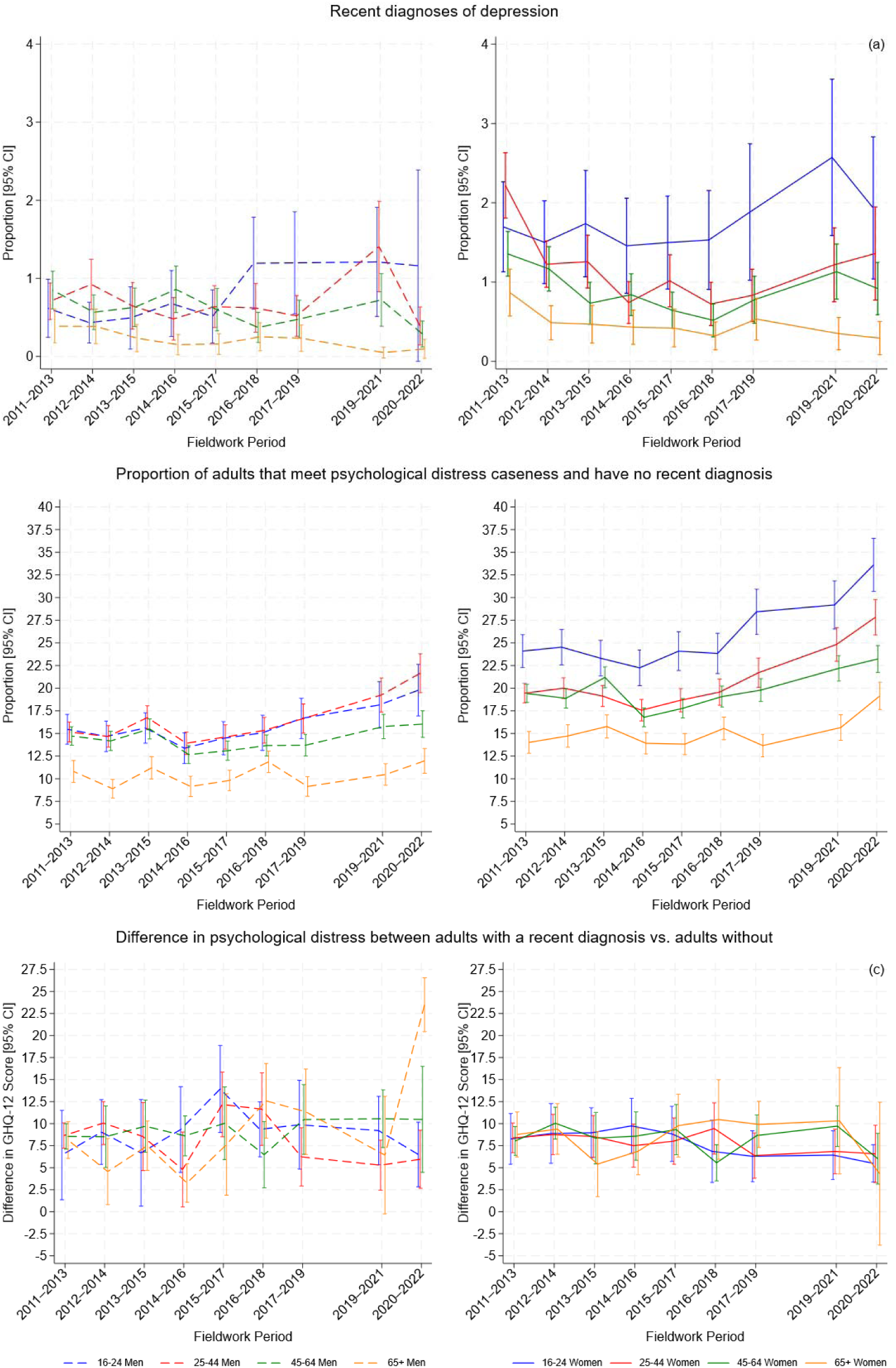
Time trends in depression diagnoses and psychological distress in the UK adult population stratified by age and sex, excluding adults from future timepoints after first reporting a diagnosis of depression. Note: Analyses of mean GHQ-12 score for adults recently diagnosed with depression are not included as there were insufficient design degrees of freedom to estimate confidence for 37 of the 72 estimates.

**Supplementary Figure 4.**
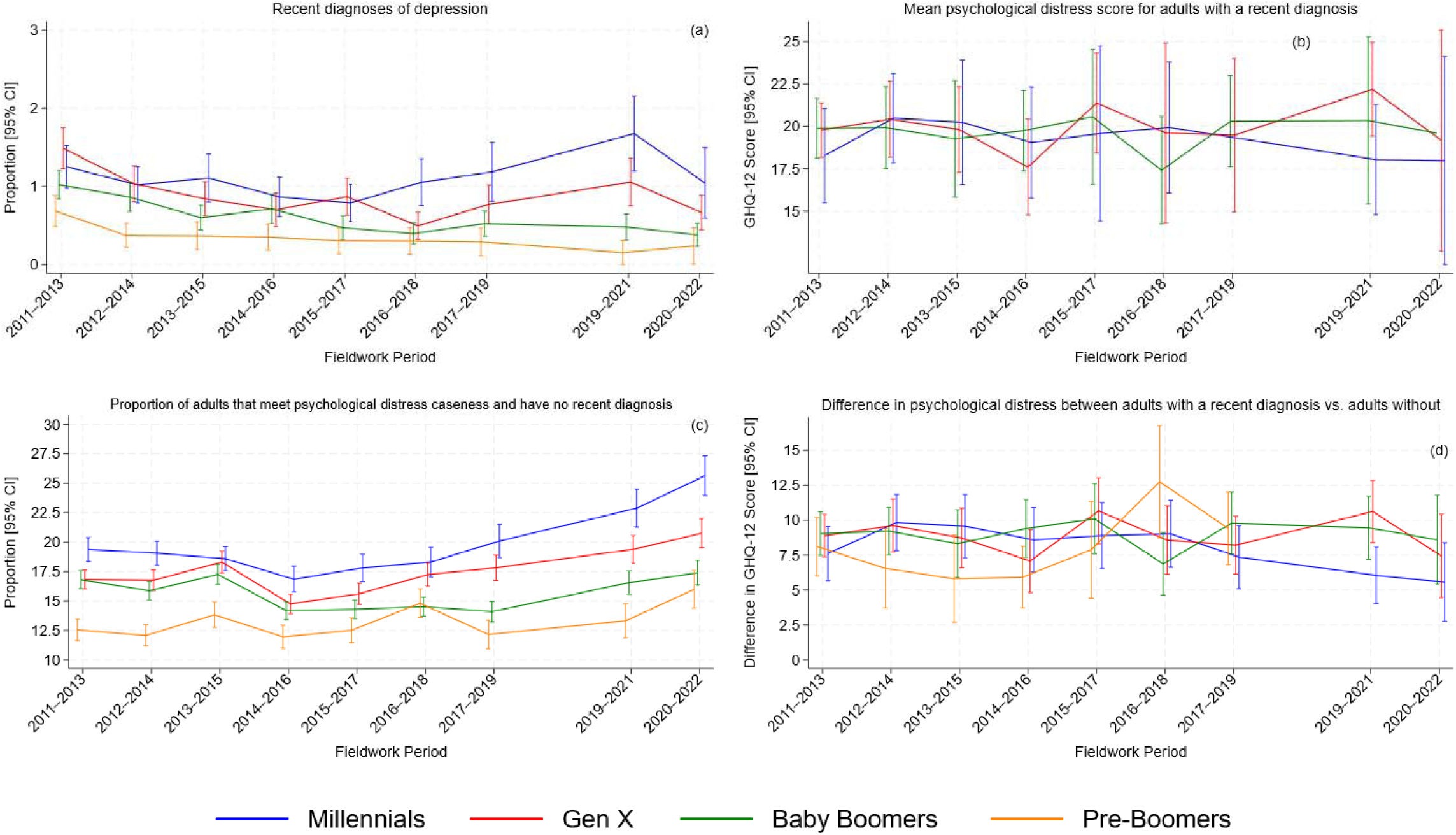
Time trends in depression diagnoses and psychological distress in the UK adult population stratified by cohort, excluding adults from future timepoints after first reporting a diagnosis of depression. Note: Analyses of mean GHQ-12 score for Pre-Boomers recently diagnosed with depression are not included in Supplementary Figure 4b as there were insufficient design degrees of freedom to estimate confidence intervals for 8 of the 9 estimates. The estimates for Baby Boomers at 2020-2022 and Millennials at 2017-2019 (95% CI: 4.34 to 33.38) excluded from Supplementary Figure 4b due to insufficient design degrees of freedom to estimate confidence intervals and large confidence intervals respectively. The estimates for Pre-Boomers at 2019-2021 (95% CI: -5.16 to 18.37) and 2020-2022 (95% CI: -3.57 to 22.84) are excluded from Supplementary Figure 4d due to large confidence intervals that reduce the readability of the figure.

**Supplementary Figure 5.**
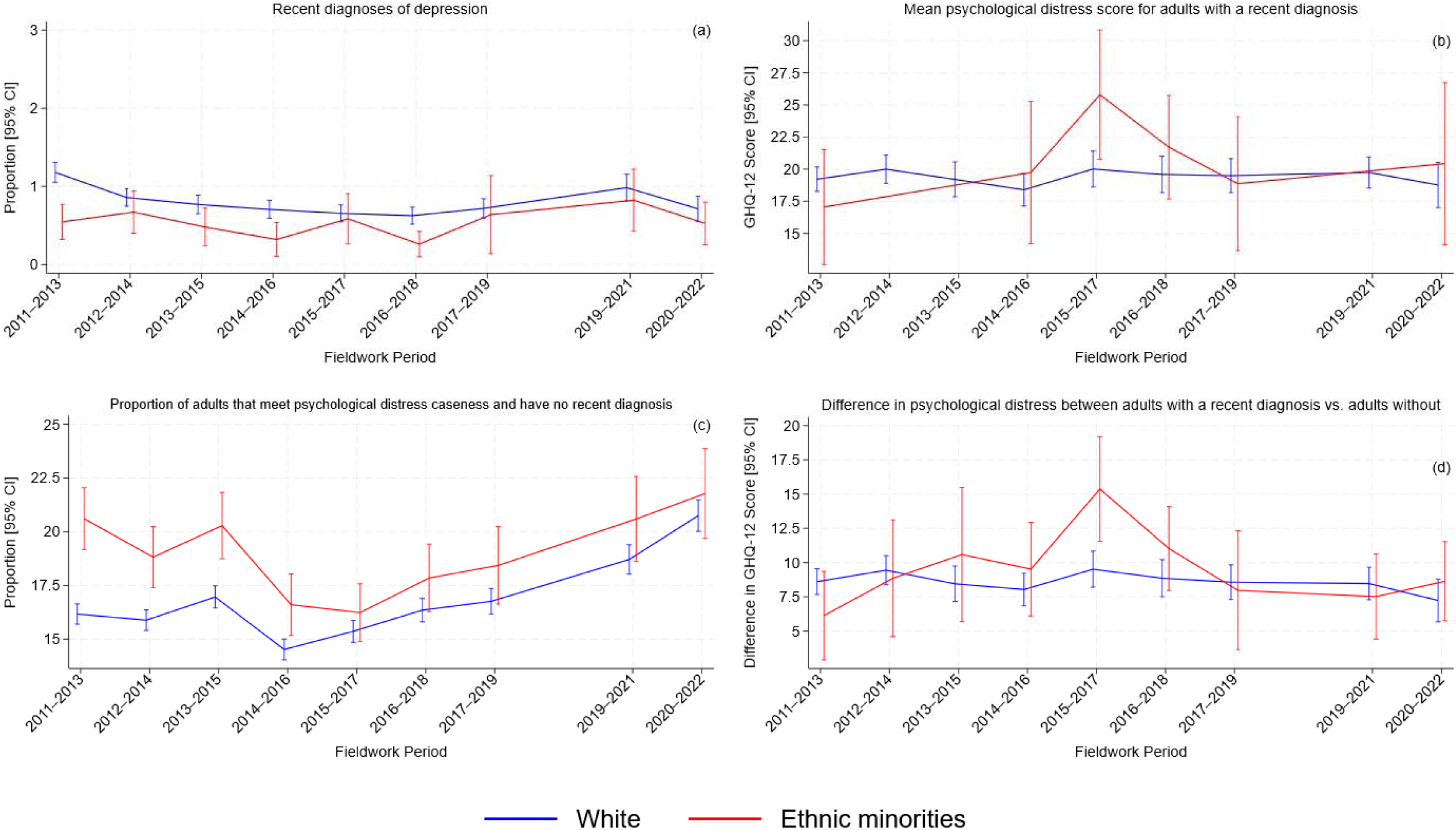
Time trends in depression diagnoses and psychological distress in the UK adult population stratified by ethnicity (binary classification), excluding adults from future timepoints after first reporting a diagnosis of depression. Note: The estimates for ethnic minorities at 2012-2014 (95% CI: -8.04 to 47.12), 2013-2015 (95% CI: -10.06 to 53.19) and 2019-2021 (95% CI: -0.97 to 39.10) are excluded from Supplementary Figure 5d due to large confidence intervals that reduce the readability of the figure.

**Supplementary Figure 6.**
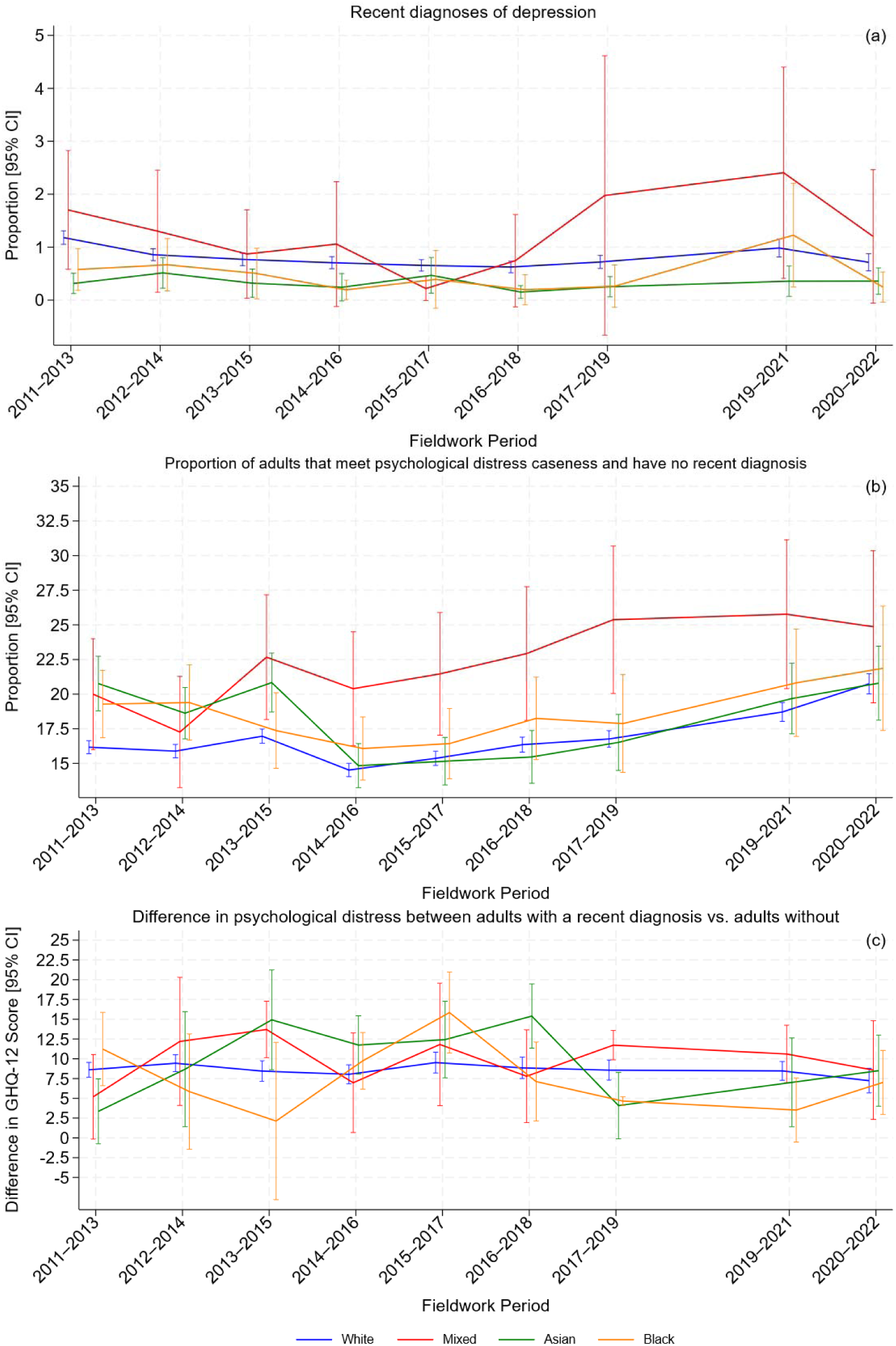
Time trends in depression diagnoses and psychological distress in the UK adult population stratified by ethnicity (four group classification), excluding adults from future timepoints after first reporting a diagnosis of depression. Note: Analyses of mean GHQ-12 score for adults recently diagnosed with depression are not included as there were insufficient design degrees of freedom to estimate confidence intervals for 18 of the 36 estimates.

**Supplementary Figure 7.**
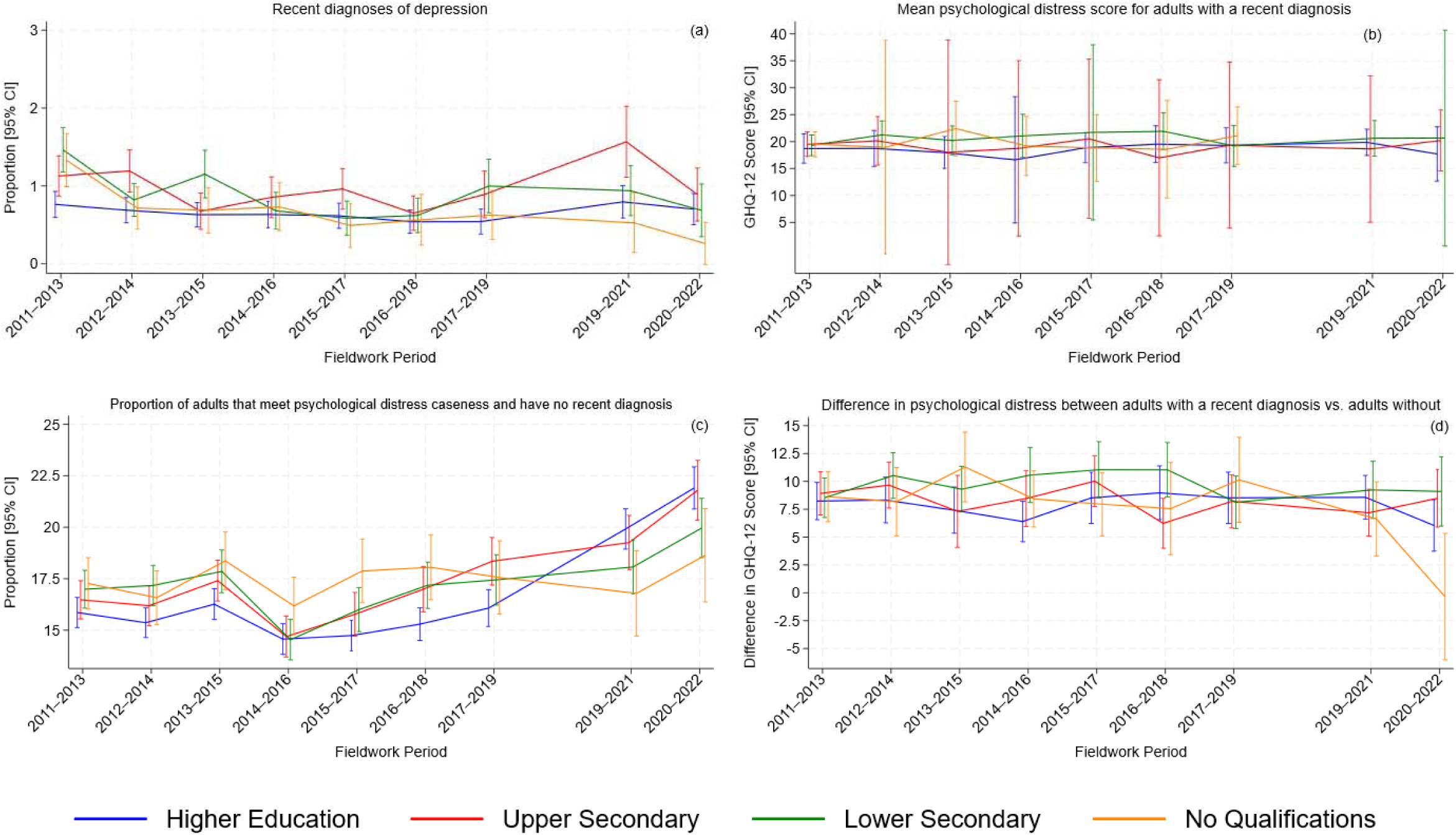
Time trends in depression diagnoses and psychological distress in the UK adult population stratified by highest educational qualification attained, excluding adults from future timepoints after first reporting a diagnosis of depression. Note: The estimates for the no qualifications group at timepoints 2019-2021 and 2020-2022 are excluded from 8b due to nsufficient design degrees of freedom to estimate confidence intervals.

**Supplementary Figure 8.**
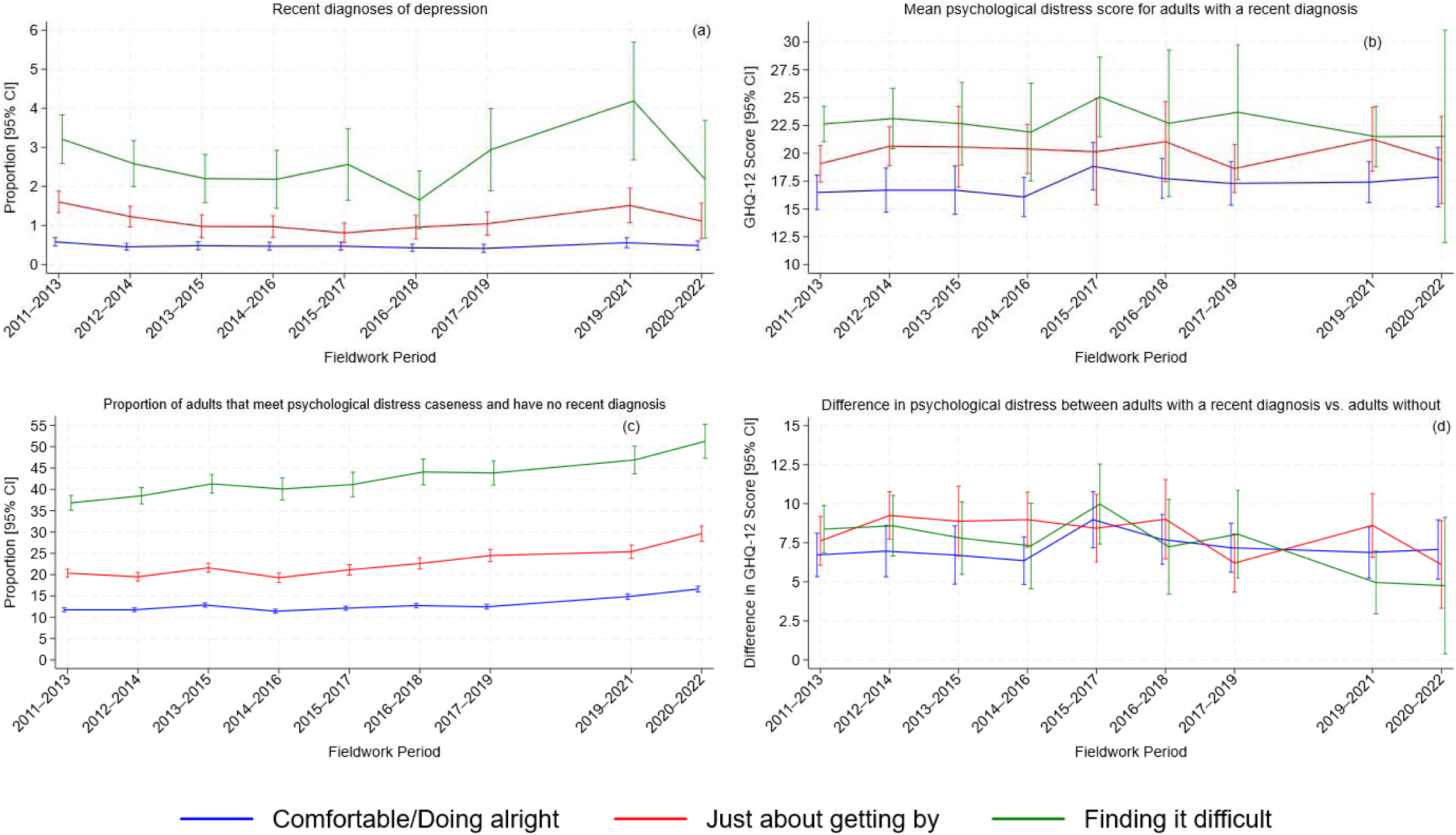
Time trends in depression diagnoses and psychological distress in the UK adult population stratified by financial stress, excluding adults from future timepoints after first reporting a diagnosis of depression.

**Supplementary Table 1.** Proportion of sample reporting a new diagnosis of depression across timepoints (%, 95% CI).

|  | Timepoint | 2011-2013 | 2012-2014 | 2013-2015 | 2014-2016 | 2015-2017 | 2016-2018 | 2017-2019 | 2019-2021 | 2020-2022 |
| --- | --- | --- | --- | --- | --- | --- | --- | --- | --- | --- |
| Caseness | No distress | 0.64 (0.54, 0.74) | 0.47 (0.39, 0.55) | 0.43 (0.34, 0.51) | 0.45 (0.35, 0.54) | 0.34 (0.26, 0.42) | 0.35 (0.27, 0.43) | 0.46 (0.36, 0.57) | 0.50 (0.38, 0.63) | 0.48 (0.36, 0.61) |
|  | Report distress | 4.96 (4.41, 5.50) | 4.36 (3.82, 4.90) | 3.93 (3.39, 4.47) | 4.10 (3.52, 4.68) | 3.53 (3.00, 4.06) | 3.54 (2.97, 4.10) | 4.26 (3.62, 4.90) | 4.59 (3.93, 5.24) | 3.75 (3.08, 4.42) |
| Sex | Men | 0.94 (0.80, 1.09) | 0.83 (0.67, 0.98) | 0.78 (0.62, 0.94) | 0.83 (0.66, 1.00) | 0.64 (0.49, 0.78) | 0.81 (0.63, 1.00) | 0.92 (0.72, 1.11) | 1.12 (0.88, 1.36) | 0.82 (0.57, 1.08) |
|  | Women | 1.93 (1.72, 2.13) | 1.55 (1.37, 1.72) | 1.44 (1.25, 1.63) | 1.29 (1.10, 1.47) | 1.17 (0.99, 1.35) | 1.08 (0.91, 1.25) | 1.49 (1.25, 1.73) | 1.64 (1.39, 1.88) | 1.61 (1.34, 1.89) |
| Age | 16-24 | 1.29 (0.90, 1.68) | 1.06 (0.76, 1.36) | 1.14 (0.76, 1.52) | 1.18 (0.81, 1.54) | 1.17 (0.82, 1.52) | 1.45 (1.01, 1.89) | 1.90 (1.29, 2.50) | 1.91 (1.30, 2.52) | 2.02 (1.24, 2.80) |
|  | 25-44 | 1.97 (1.70, 2.24) | 1.58 (1.33, 1.83) | 1.49 (1.20, 1.77) | 1.04 (0.80, 1.28) | 1.15 (0.91, 1.39) | 1.13 (0.86, 1.40) | 1.14 (0.85, 1.43) | 1.72 (1.32, 2.12) | 1.51 (1.08, 1.93) |
|  | 45-64 | 1.51 (1.30, 1.72) | 1.27 (1.07, 1.48) | 1.20 (0.98, 1.42) | 1.37 (1.13, 1.61) | 0.96 (0.76, 1.17) | 0.99 (0.77, 1.21) | 1.34 (1.07, 1.62) | 1.67 (1.34, 2.00) | 1.42 (1.10, 1.73) |
|  | 65+ | 0.74 (0.55, 0.93) | 0.67 (0.49, 0.85) | 0.54 (0.37, 0.71) | 0.60 (0.42, 0.78) | 0.44 (0.28, 0.60) | 0.43 (0.27, 0.58) | 0.75 (0.53, 0.97) | 0.46 (0.30, 0.62) | 0.40 (0.24, 0.56) |
| Age and sex | Male 16-24 | 0.71 (0.29, 1.13) | 0.48 (0.21, 0.76) | 0.55 (0.15, 0.95) | 0.79 (0.33, 1.25) | 0.75 (0.34, 1.16) | 1.30 (0.67, 1.92) | 1.42 (0.73, 2.10) | 1.19 (0.50, 1.87) | 1.34 (0.12, 2.56) |
|  | Female 16-24 | 1.91 (1.26, 2.56) | 1.71 (1.15, 2.26) | 1.80 (1.14, 2.46) | 1.61 (1.00, 2.22) | 1.63 (1.04, 2.21) | 1.62 (1.00, 2.24) | 2.38 (1.38, 3.37) | 2.59 (1.61, 3.57) | 2.63 (1.61, 3.65) |
|  | Male 25-44 | 0.93 (0.66, 1.19) | 1.24 (0.89, 1.59) | 0.99 (0.64, 1.35) | 0.73 (0.43, 1.03) | 0.83 (0.52, 1.14) | 0.95 (0.57, 1.33) | 0.87 (0.47, 1.27) | 1.61 (1.02, 2.21) | 0.96 (0.49, 1.44) |
|  | Female 25-44 | 2.89 (2.43, 3.35) | 1.88 (1.53, 2.24) | 1.93 (1.54, 2.31) | 1.32 (0.97, 1.67) | 1.43 (1.06, 1.80) | 1.29 (0.91, 1.67) | 1.38 (0.97, 1.78) | 1.81 (1.29, 2.32) | 1.99 (1.33, 2.64) |
|  | Male 45-64 | 1.35 (1.05, 1.65) | 0.84 (0.59, 1.10) | 1.00 (0.70, 1.31) | 1.20 (0.86, 1.54) | 0.71 (0.43, 0.98) | 0.83 (0.51, 1.16) | 1.01 (0.67, 1.35) | 1.41 (0.95, 1.87) | 1.01 (0.60, 1.43) |
|  | Female 45-64 | 1.65 (1.36, 1.95) | 1.67 (1.35, 1.98) | 1.38 (1.06, 1.69) | 1.52 (1.19, 1.86) | 1.20 (0.90, 1.49) | 1.14 (0.85, 1.43) | 1.65 (1.25, 2.05) | 1.91 (1.46, 2.36) | 1.79 (1.33, 2.24) |
|  | Male 65+ | 0.50 (0.27, 0.74) | 0.50 (0.26, 0.74) | 0.36 (0.16, 0.56) | 0.44 (0.21, 0.67) | 0.25 (0.09, 0.40) | 0.34 (0.14, 0.55) | 0.55 (0.28, 0.81) | 0.24 (0.07, 0.41) | 0.19 (0.02, 0.37) |
|  | Female 65+ | 0.96 (0.66, 1.26) | 0.82 (0.56, 1.09) | 0.72 (0.44, 0.99) | 0.75 (0.47, 1.02) | 0.61 (0.34, 0.88) | 0.50 (0.28, 0.72) | 0.93 (0.60, 1.27) | 0.65 (0.39, 0.91) | 0.58 (0.32, 0.84) |
| Cohort | Pre-Boomers | 0.76 (0.56, 0.97) | 0.59 (0.41, 0.77) | 0.51 (0.31, 0.70) | 0.56 (0.35, 0.77) | 0.42 (0.23, 0.61) | 0.38 (0.18, 0.57) | 0.67 (0.39, 0.96) | 0.26 (0.08, 0.44) | 0.26 (0.03, 0.49) |
|  | Baby Boomers | 1.41 (1.20, 1.62) | 1.19 (0.98, 1.39) | 1.05 (0.85, 1.26) | 1.26 (1.03, 1.49) | 0.79 (0.60, 0.98) | 0.80 (0.61, 0.99) | 1.12 (0.87, 1.36) | 1.07 (0.82, 1.32) | 0.94 (0.69, 1.20) |
|  | Generation X | 1.89 (1.60, 2.17) | 1.61 (1.33, 1.89) | 1.39 (1.12, 1.67) | 1.09 (0.84, 1.35) | 1.14 (0.89, 1.39) | 1.01 (0.76, 1.27) | 1.41 (1.08, 1.75) | 1.79 (1.41, 2.18) | 1.38 (1.04, 1.71) |
|  | Millennials | 1.62 (1.31, 1.93) | 1.29 (1.03, 1.55) | 1.41 (1.08, 1.74) | 1.23 (0.94, 1.52) | 1.11 (0.83, 1.39) | 1.45 (1.10, 1.81) | 1.57 (1.16, 1.99) | 1.87 (1.39, 2.34) | 1.70 (1.17, 2.24) |
| Ethnicity | White | 1.51 (1.37, 1.65) | 1.22 (1.09, 1.35) | 1.16 (1.02, 1.30) | 1.12 (0.98, 1.26) | 0.94 (0.81, 1.06) | 0.99 (0.86, 1.13) | 1.23 (1.07, 1.39) | 1.42 (1.23, 1.62) | 1.28 (1.08, 1.49) |
|  | Ethnic minorities | 0.84 (0.51, 1.16) | 1.03 (0.70, 1.35) | 0.75 (0.47, 1.04) | 0.43 (0.20, 0.66) | 0.67 (0.35, 0.99) | 0.45 (0.24, 0.65) | 0.98 (0.46, 1.50) | 1.10 (0.67, 1.52) | 0.85 (0.38, 1.31) |
|  | Mixed | 2.37 (0.79, 3.96) | 1.52 (0.38, 2.66) | 1.70 (0.53, 2.86) | 0.98 (-0.11, 2.06) | 0.53 (0.04, 1.01) | 1.04 (0.09, 1.98) | 2.48 (-0.03, 5.00) | 2.81 (0.85, 4.77) | 1.51 (0.28, 2.75) |
|  | Asian | 0.54 (0.19, 0.88) | 0.88 (0.50, 1.26) | 0.54 (0.23, 0.86) | 0.44 (0.14, 0.75) | 0.56 (0.21, 0.92) | 0.35 (0.15, 0.56) | 0.43 (0.18, 0.67) | 0.51 (0.18, 0.84) | 0.41 (0.16, 0.66) |
|  | Black | 0.78 (0.26, 1.29) | 1.08 (0.41, 1.76) | 0.63 (0.12, 1.13) | 0.19 (0.01, 0.36) | 0.39 (-0.13, 0.91) | 0.23 (-0.05, 0.51) | 0.45 (-0.00, 0.90) | 1.38 (0.39, 2.36) | 1.10 (-0.49, 2.69) |
| Education | Higher Education | 1.02 (0.83, 1.21) | 0.97 (0.79, 1.16) | 0.94 (0.76, 1.13) | 0.89 (0.70, 1.08) | 0.81 (0.63, 0.99) | 0.87 (0.67, 1.06) | 1.06 (0.84, 1.29) | 1.20 (0.96, 1.45) | 1.19 (0.94, 1.44) |
|  | Upper Secondary | 1.51 (1.21, 1.81) | 1.48 (1.19, 1.76) | 1.20 (0.92, 1.49) | 1.16 (0.87, 1.45) | 1.16 (0.89, 1.43) | 0.89 (0.65, 1.14) | 1.23 (0.90, 1.56) | 1.94 (1.45, 2.42) | 1.49 (1.06, 1.92) |
|  | Lower Secondary | 1.82 (1.51, 2.13) | 1.24 (0.99, 1.49) | 1.38 (1.07, 1.70) | 1.17 (0.88, 1.46) | 0.99 (0.72, 1.27) | 1.15 (0.84, 1.46) | 1.68 (1.25, 2.12) | 1.41 (1.02, 1.80) | 1.26 (0.83, 1.70) |
|  | No Qualifications | 1.64 (1.26, 2.01) | 1.17 (0.85, 1.50) | 1.18 (0.82, 1.54) | 1.40 (0.97, 1.83) | 0.79 (0.47, 1.12) | 0.92 (0.52, 1.32) | 1.10 (0.69, 1.50) | 0.84 (0.43, 1.25) | 0.67 (0.24, 1.09) |
| Financial stress | Low | 0.77 (0.64, 0.89) | 0.66 (0.55, 0.76) | 0.69 (0.57, 0.81) | 0.69 (0.56, 0.81) | 0.64 (0.52, 0.76) | 0.62 (0.51, 0.74) | 0.70 (0.57, 0.84) | 0.82 (0.68, 0.97) | 0.76 (0.62, 0.91) |
|  | Moderate | 2.10 (1.79, 2.41) | 1.64 (1.35, 1.93) | 1.35 (1.05, 1.65) | 1.54 (1.22, 1.86) | 1.33 (1.02, 1.64) | 1.53 (1.15, 1.91) | 1.90 (1.50, 2.30) | 2.02 (1.55, 2.50) | 2.19 (1.61, 2.78) |
|  | High | 3.79 (3.18, 4.41) | 3.61 (2.96, 4.27) | 3.69 (2.97, 4.40) | 3.65 (2.79, 4.50) | 2.88 (2.04, 3.73) | 2.91 (2.07, 3.75) | 4.12 (3.05, 5.19) | 5.47 (3.99, 6.94) | 3.99 (2.33, 5.65) |

**Supplementary Table 2.** Mean GHQ-12 score for adults reporting a new diagnosis of depression across timepoints (mean, 95% CI).

|  | Timepoint | 2011-2013 | 2012-2014 | 2013-2015 | 2014-2016 | 2015-2017 | 2016-2018 | 2017-2019 | 2019-2021 | 2020-2022 |
| --- | --- | --- | --- | --- | --- | --- | --- | --- | --- | --- |
|  | Whole sample | 19.28 (18.52, 20.04) | 20.51 (19.61, 21.41) | 19.97 (19.03, 20.91) | 19.41 (18.46, 20.36) | 20.60 (19.55, 21.66) | 19.92 (18.85, 21.00) | 20.28 (19.29, 21.28) | 20.31 (19.38, 21.25) | 20.74 (19.51, 21.97) |
| Sex | Men | 18.88 (17.57, 20.19) | 19.68 (17.98, 21.38) | 20.00 (18.16, 21.85) | 18.13 (16.41, 19.85) | 21.21 (19.02, 23.40) | 19.92 (17.71, 22.13) | 20.03 (17.99, 22.08) | 19.56 (17.71, 21.41) | 19.96 (17.62, 22.30) |
|  | Women | 19.46 (18.50, 20.41) | 20.94 (19.88, 22.00) | 19.95 (18.79, 21.10) | 20.19 (18.99, 21.39) | 20.30 (19.00, 21.60) | 19.92 (18.53, 21.32) | 20.43 (19.19, 21.67) | 20.79 (19.56, 22.02) | 21.10 (19.51, 22.69) |
| Age | 16-24 | 18.61 (13.36, 23.87) | 20.73 (16.99, 24.46) | 18.92 (1.75, 36.08) | 19.88 (16.00, 23.77) | 20.49 (17.34, 23.64) | 18.35 (13.04, 23.65) | 19.36 (14.29, 24.43) | 19.79 (17.02, 22.56) | 19.84 (15.09, 24.60) |
|  | 25-44 | 19.70 (18.46, 20.93) | 20.82 (19.33, 22.31) | 20.03 (18.18, 21.88) | 19.55 (17.29, 21.82) | 21.01 (18.92, 23.09) | 20.85 (18.37, 23.32) | 19.55 (16.81, 22.29) | 18.47 (16.55, 20.39) | 20.43 (17.63, 23.22) |
|  | 45-64 | 19.40 (18.22, 20.58) | 21.26 (19.77, 22.76) | 20.91 (19.33, 22.49) | 20.08 (18.61, 21.54) | 20.65 (18.63, 22.66) | 20.18 (18.41, 21.96) | 21.29 (19.68, 22.90) | 21.82 (20.20, 23.44) | 21.27 (19.03, 23.51) |
|  | 65+ | 18.06 (15.73, 20.40) | 17.27 (13.97, 20.58) | 18.05 | 16.46 | 19.37 (3.95, 34.79) | 19.32 (14.33, 24.31) | 20.32 (10.70, 29.94) | 20.55 | 21.62 |
| Age and sex | Male 16-24 | 16.22 | 20.63 | 15.97 | 19.06 | 21.75 (-6.31, 49.82) | 18.05 | 20.53 (-6.11, 47.17) | 20.23 | 18.52 |
|  | Female 16-24 | 19.37 (2.74, 36.00) | 20.75 (15.78, 25.73) | 19.93 (1.97, 37.90) | 20.34 (13.96, 26.71) | 19.86 (1.34, 38.38) | 18.60 (11.62, 25.57) | 18.64 (1.46, 35.81) | 19.60 (15.78, 23.41) | 20.44 (4.44, 36.44) |
|  | Male 25-44 | 18.98 (16.40, 21.57) | 20.38 (17.35, 23.41) | 21.53 (15.12, 27.93) | 17.06 | 21.89 (0.72, 43.07) | 20.70 | 17.85 | 16.82 | 18.21 |
|  | Female 25-44 | 19.90 (18.39, 21.41) | 21.10 (19.03, 23.16) | 19.39 (17.17, 21.60) | 20.81 (18.21, 23.41) | 20.55 (17.75, 23.35) | 20.94 (17.18, 24.70) | 20.51 (16.13, 24.89) | 19.80 (17.46, 22.14) | 21.38 (16.60, 26.16) |
|  | Male 45-64 | 19.80 (17.64, 21.97) | 19.95 (2.74, 37.17) | 20.53 (17.40, 23.67) | 19.23 (16.78, 21.68) | 20.83 (15.46, 26.20) | 20.61 (14.49, 26.73) | 21.20 (16.85, 25.55) | 21.62 (16.84, 26.41) | 20.73 (1.76, 39.70) |
|  | Female 45-64 | 19.08 (17.52, 20.64) | 21.86 (20.13, 23.60) | 21.16 (18.24, 24.08) | 20.71 (18.52, 22.91) | 20.55 (17.86, 23.24) | 19.90 (17.58, 22.21) | 21.35 (19.39, 23.30) | 21.95 (19.63, 24.27) | 21.55 (17.69, 25.42) |
|  | Male 65+ | 17.02 (-2.69, 36.74) | 16.26 | 17.04 | 15.00 | 18.96 | 19.46 | 20.03 | 18.41 | 27.62 |
|  | Female 65+ | 18.57 (3.66, 33.48) | 17.87 (12.75, 22.99) | 18.53 | 17.24 | 19.52 | 19.23 (12.82, 25.65) | 20.46 | 21.25 | 19.82 |
| Cohort | Pre-Boomers | 18.07 (15.66, 20.48) | 16.42 (12.76, 20.08) | 17.75 | 16.00 | 17.00 | 21.92 | 20.53 | 17.43 | 20.83 |
|  | Baby Boomers | 19.78 (18.49, 21.08) | 20.10 (18.60, 21.60) | 20.18 (18.32, 22.05) | 19.97 (18.33, 21.60) | 20.97 (18.85, 23.09) | 18.97 (16.78, 21.15) | 20.76 (18.83, 22.69) | 21.25 (18.85, 23.65) | 21.33 |
|  | Generation X | 19.31 (18.03, 20.60) | 21.81 (20.16, 23.46) | 21.23 (19.40, 23.05) | 19.54 (17.22, 21.86) | 22.10 (19.90, 24.30) | 20.95 (18.33, 23.57) | 20.86 (18.66, 23.05) | 21.97 (20.13, 23.81) | 21.86 (18.86, 24.85) |
|  | Millennials | 19.16 (17.13, 21.20) | 20.83 (18.55, 23.11) | 19.17 (16.40, 21.94) | 19.94 (17.64, 22.25) | 19.31 (16.42, 22.20) | 19.60 (17.02, 22.19) | 19.47 (16.90, 22.04) | 18.26 (15.75, 20.76) | 19.36 (16.39, 22.32) |
| Ethnicity | White | 19.34 (18.56, 20.11) | 20.61 (19.68, 21.53) | 19.80 (18.82, 20.77) | 19.39 (18.40, 20.38) | 20.34 (19.24, 21.45) | 19.95 (18.84, 21.07) | 20.25 (19.20, 21.31) | 20.39 (19.40, 21.38) | 20.47 (19.22, 21.72) |
|  | Ethnic minorities | 17.00 (13.63, 20.37) | 19.63 (14.44, 24.81) | 22.59 (15.28, 29.89) | 19.48 (14.75, 24.22) | 24.82 (20.65, 28.99) | 21.18 (17.70, 24.66) | 20.72 (17.18, 24.25) | 19.39 (16.01, 22.76) | 24.47 (18.07, 30.87) |
|  | Mixed | 14.35 | 22.26 | 23.98 | 17.84 (-22.73, 58.42) | 16.43 (3.18, 29.67) | 19.59 | 22.69 | 21.72 | 23.41 (-9.10, 55.92) |
|  | Asian | 16.52 (10.95, 22.10) | 21.10 (13.65, 28.54) | 26.15 | 20.28 (-1.98, 42.55) | 23.33 (18.03, 28.63) | 24.69 (17.98, 31.39) | 19.30 (9.62, 28.98) | 19.76 (-7.43, 46.95) | 20.34 (-4.88, 45.56) |
|  | Black | 19.97 | 14.27 | 14.71 (-38.88, 68.29) | 19.52 | 25.30 | 17.44 (-8.08, 42.96) | 19.89 | 14.68 | 30.93 (-15.75, 77.60) |
| Education | Higher Education | 18.82 (17.06, 20.58) | 19.03 (16.76, 21.30) | 18.22 (16.11, 20.32) | 17.58 (15.01, 20.16) | 19.40 (17.31, 21.49) | 19.61 (17.05, 22.17) | 19.24 (17.19, 21.30) | 19.80 (18.04, 21.56) | 19.75 (17.38, 22.11) |
|  | Upper Secondary | 19.11 (17.27, 20.96) | 20.15 (17.62, 22.67) | 19.68 (16.82, 22.54) | 19.00 (14.61, 23.38) | 19.81 (15.56, 24.06) | 17.50 (14.54, 20.47) | 19.03 (7.19, 30.88) | 20.01 (16.70, 23.31) | 21.46 (16.54, 26.38) |
|  | Lower Secondary | 19.29 (17.80, 20.78) | 21.80 (19.88, 23.71) | 20.23 (18.06, 22.40) | 20.72 (18.33, 23.10) | 22.40 (19.71, 25.08) | 21.76 (19.59, 23.92) | 20.52 (18.17, 22.86) | 20.63 (18.33, 22.93) | 21.26 (16.65, 25.87) |
|  | No Qualifications | 20.42 (18.40, 22.44) | 20.89 (18.08, 23.70) | 22.24 (19.14, 25.35) | 21.25 (18.29, 24.21) | 20.06 (15.31, 24.82) | 20.73 (12.51, 28.94) | 22.91 (19.24, 26.57) | 19.23 (1.37, 37.10) | 22.85 |
| Financial stress | Low | 17.24 (15.89, 18.59) | 16.69 (15.24, 18.14) | 16.73 (15.14, 18.33) | 16.72 (15.40, 18.04) | 18.79 (17.16, 20.43) | 17.41 (15.93, 18.88) | 17.40 (15.96, 18.84) | 17.72 (16.27, 19.17) | 18.73 (16.96, 20.51) |
|  | Moderate | 18.78 (17.51, 20.05) | 20.83 (19.32, 22.34) | 20.50 (18.51, 22.50) | 20.47 (18.78, 22.16) | 20.52 (18.24, 22.81) | 20.80 (18.56, 23.03) | 20.66 (18.91, 22.40) | 22.02 (20.15, 23.90) | 21.53 (18.87, 24.19) |
|  | High | 22.42 (21.05, 23.79) | 24.60 (23.04, 26.15) | 23.80 (21.88, 25.71) | 23.35 (20.97, 25.73) | 25.60 (23.15, 28.05) | 24.54 (21.46, 27.62) | 24.61 (22.13, 27.08) | 22.56 (20.56, 24.56) | 24.00 (19.65, 28.34) |

**Supplementary Table 3.**
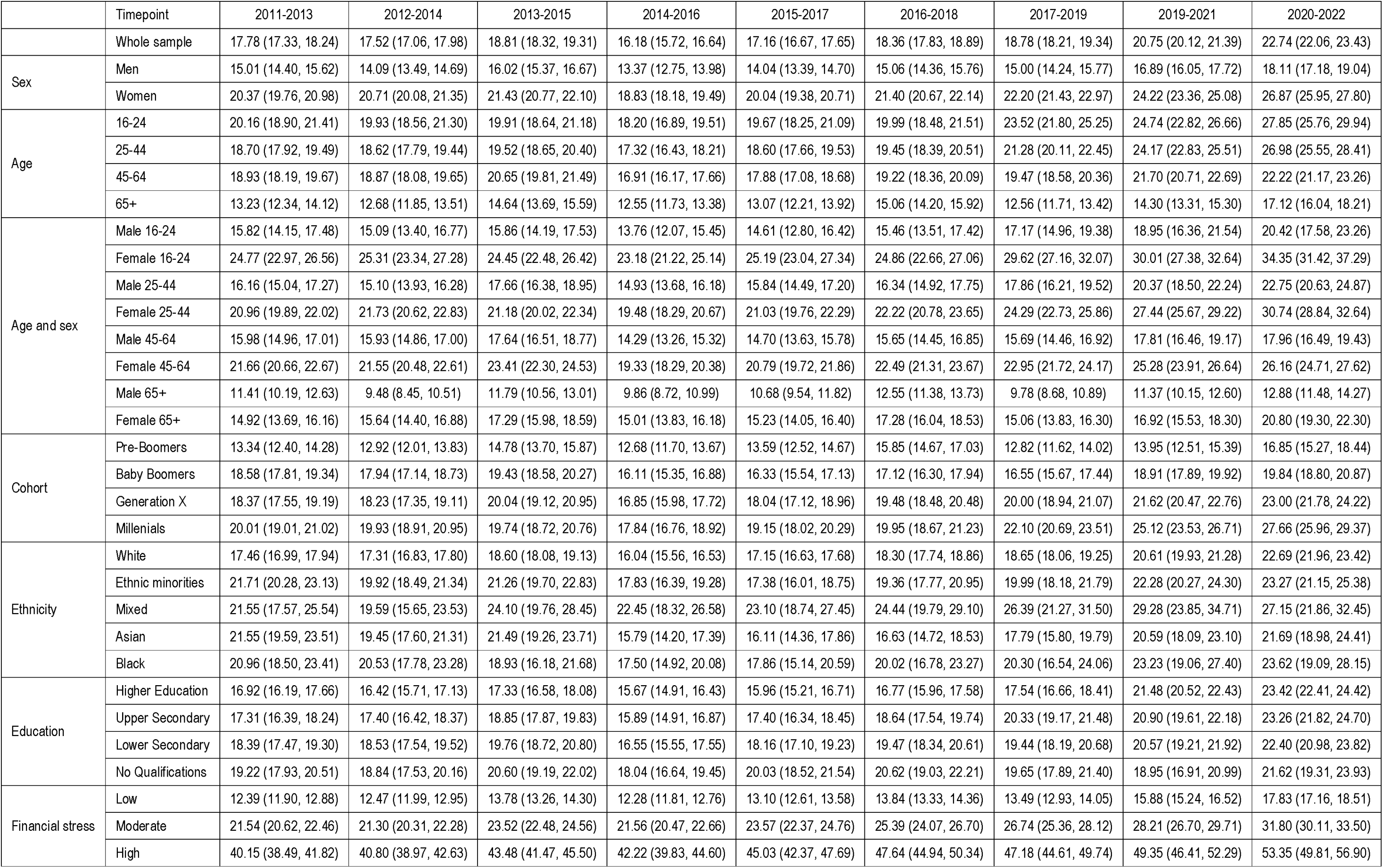
Proportion of sample reporting psychological distress without a recent depression diagnosis across timepoints (%, 95% CI).

**Supplementary Table 4.** Mean difference in psychological distress (GHQ-12 score) between adults with a new diagnosis of depression and adults without a new diagnosis of depression across timepoints (*b*, 95% CI).

|  | Timepoint | 2011-2013 | 2012-2014 | 2013-2015 | 2014-2016 | 2015-2017 | 2016-2018 | 2017-2019 | 2019-2021 | 2020-2022 |
| --- | --- | --- | --- | --- | --- | --- | --- | --- | --- | --- |
|  | Whole sample | 8.39 (7.63, 9.14) | 9.67 (8.78, 10.55) | 8.90 (7.98, 9.82) | 8.78 (7.85, 9.70) | 9.78 (8.75, 10.81) | 8.83 (7.77, 9.89) | 8.99 (8.02, 9.96) | 8.67 (7.74, 9.60) | 8.80 (7.61, 9.98) |
| Sex | Men | 8.57 (7.32, 9.82) | 9.50 (7.97, 11.04) | 9.52 (7.87, 11.18) | 8.03 (6.51, 9.56) | 10.97 (9.08, 12.85) | 9.42 (7.73, 11.11) | 9.34 (7.64, 11.05) | 8.54 (6.93, 10.15) | 8.80 (6.92, 10.69) |
|  | Women | 8.02 (7.08, 8.96) | 9.47 (8.44, 10.50) | 8.33 (7.21, 9.45) | 9.05 (7.90, 10.20) | 8.93 (7.69, 10.16) | 8.29 (6.94, 9.64) | 8.59 (7.40, 9.77) | 8.58 (7.38, 9.77) | 8.45 (6.95, 9.95) |
| Age | 16-24 | 7.92 (5.51, 10.32) | 10.22 (7.56, 12.87) | 8.28 (5.64, 10.92) | 9.36 (6.98, 11.75) | 9.82 (7.43, 12.21) | 7.37 (4.95, 9.80) | 7.69 (5.36, 10.01) | 7.68 (5.46, 9.90) | 7.26 (5.06, 9.46) |
|  | 25-44 | 8.58 (7.39, 9.76) | 9.69 (8.29, 11.10) | 8.78 (7.11, 10.45) | 8.72 (6.83, 10.62) | 9.88 (7.98, 11.77) | 9.55 (7.53, 11.56) | 7.76 (5.57, 9.95) | 6.22 (4.62, 7.82) | 7.70 (5.45, 9.94) |
|  | 45-64 | 8.13 (7.00, 9.25) | 9.99 (8.61, 11.37) | 9.34 (7.94, 10.74) | 9.18 (7.80, 10.55) | 9.48 (7.69, 11.27) | 8.72 (7.10, 10.34) | 9.73 (8.26, 11.20) | 9.91 (8.42, 11.39) | 9.23 (7.36, 11.09) |
|  | 65+ | 7.93 (6.03, 9.84) | 7.22 (5.20, 9.25) | 7.68 (5.26, 10.09) | 6.36 (4.58, 8.15) | 9.29 (6.90, 11.68) | 8.89 (6.62, 11.17) | 10.14 (8.64, 11.63) | 10.04 (6.86, 13.22) | 10.83 (6.50, 15.15) |
| Age and sex | Male 16-24 | 6.36 (1.29, 11.44) | 11.02 (5.90, 16.14) | 6.24 (0.90, 11.58) | 9.40 (5.24, 13.55) | 11.96 (7.78, 16.15) | 8.02 (4.34, 11.70) | 9.88 (5.73, 14.03) | 9.06 (5.18, 12.95) | 7.39 (3.68, 11.11) |
|  | Female 16-24 | 7.77 (5.20, 10.35) | 9.23 (6.15, 12.30) | 8.28 (5.52, 11.03) | 8.85 (5.95, 11.75) | 8.21 (5.31, 11.10) | 6.61 (3.42, 9.81) | 5.97 (3.30, 8.65) | 6.63 (3.93, 9.32) | 6.55 (4.05, 9.06) |
|  | Male 25-44 | 8.58 (7.39, 9.76) | 9.89 (7.74, 12.04) | 10.72 (7.81, 13.63) | 6.63 (3.09, 10.17) | 11.28 (8.00, 14.56) | 9.86 (6.82, 12.91) | 6.57 (3.00, 10.14) | 5.21 (2.65, 7.78) | 6.22 (3.57, 8.87) |
|  | Female 25-44 | 8.25 (6.85, 9.65) | 9.40 (7.58, 11.21) | 7.74 (5.85, 9.63) | 9.61 (7.51, 11.71) | 8.96 (6.64, 11.29) | 9.23 (6.56, 11.90) | 8.26 (5.57, 10.96) | 6.98 (5.10, 8.86) | 7.99 (5.02, 10.95) |
|  | Male 45-64 | 8.13 (7.00, 9.25) | 9.26 (6.59, 11.92) | 9.53 (7.26, 11.80) | 8.86 (6.91, 10.82) | 10.25 (6.84, 13.65) | 9.76 (6.96, 12.56) | 10.20 (7.51, 12.89) | 10.32 (8.12, 12.53) | 9.32 (6.38, 12.26) |
|  | Female 45-64 | 7.31 (5.87, 8.76) | 10.05 (8.50, 11.60) | 9.08 (7.27, 10.88) | 9.32 (7.39, 11.25) | 8.84 (6.78, 10.90) | 7.88 (5.91, 9.85) | 9.26 (7.58, 10.94) | 9.48 (7.50, 11.45) | 8.92 (6.53, 11.31) |
|  | Male 65+ | 7.93 (6.03, 9.84) | 6.84 (2.98, 10.70) | 7.21 (5.23, 9.19) | 5.39 (2.03, 8.75) | 9.36 (4.96, 13.77) | 9.53 (4.96, 14.10) | 10.38 (7.49, 13.27) | 8.37 (3.68, 13.06) | 17.56 (11.78, 23.34) |
|  | Female 65+ | 8.02 (5.66, 10.38) | 7.24 (4.93, 9.55) | 7.65 (4.20, 11.09) | 6.70 (4.64, 8.76) | 9.01 (6.15, 11.86) | 8.37 (5.44, 11.29) | 9.80 (7.75, 11.86) | 10.32 (6.38, 14.26) | 8.39 (3.33, 13.44) |
| Cohort | Pre-Boomers | 7.93 (5.96, 9.89) | 6.35 (4.10, 8.59) | 7.36 (4.28, 10.44) | 5.89 (4.11, 7.68) | 6.90 (4.15, 9.64) | 11.44 (8.67, 14.20) | 10.32 (8.47, 12.18) | 7.04 (-0.27, 14.34) | 10.19 (-1.28, 21.66) |
|  | Baby Boomers | 8.59 (7.37, 9.80) | 8.99 (7.62, 10.36) | 8.84 (7.22, 10.46) | 9.25 (7.73, 10.77) | 10.13 (8.35, 11.90) | 7.94 (6.08, 9.80) | 9.78 (8.18, 11.38) | 9.94 (8.10, 11.78) | 9.86 (7.43, 12.29) |
|  | Generation X | 8.13 (6.90, 9.37) | 10.68 (9.14, 12.22) | 9.80 (8.15, 11.44) | 8.63 (6.70, 10.56) | 10.91 (8.93, 12.89) | 9.47 (7.60, 11.33) | 9.17 (7.35, 10.98) | 9.97 (8.31, 11.63) | 9.66 (7.53, 11.79) |
|  | Millennials | 8.35 (6.64, 10.06) | 10.01 (8.17, 11.84) | 8.29 (6.33, 10.24) | 9.29 (7.43, 11.14) | 8.35 (6.31, 10.40) | 8.39 (6.21, 10.56) | 7.60 (5.54, 9.66) | 5.88 (4.11, 7.66) | 6.54 (4.16, 8.93) |
| Ethnicity | White | 8.46 (7.70, 9.23) | 9.76 (8.85, 10.67) | 8.73 (7.78, 9.69) | 8.73 (7.78, 9.69) | 9.50 (8.44, 10.57) | 8.85 (7.75, 9.94) | 8.95 (7.93, 9.98) | 8.76 (7.77, 9.74) | 8.54 (7.36, 9.73) |
|  | Ethnic minorities | 5.88 (3.27, 8.48) | 8.71 (5.51, 11.92) | 11.42 (8.08, 14.77) | 9.04 (6.12, 11.96) | 14.16 (10.62, 17.70) | 10.25 (7.46, 13.04) | 9.56 (6.57, 12.54) | 7.59 (5.18, 9.99) | 12.41 (7.20, 17.63) |
|  | Mixed | 3.19 (-1.07, 7.46) | 11.35 (4.21, 18.49) | 12.25 (8.91, 15.59) | 6.56 (0.26, 12.86) | 4.53 (-1.58, 10.64) | 7.55 (3.58, 11.51) | 10.64 (8.64, 12.64) | 9.02 (5.75, 12.29) | 10.31 (5.17, 15.44) |
|  | Asian | 5.34 (1.93, 8.76) | 10.05 (5.44, 14.65) | 14.93 (10.54, 19.33) | 10.00 (6.56, 13.44) | 12.81 (8.76, 16.85) | 14.06 (9.90, 18.22) | 8.28 (3.83, 12.73) | 7.99 (3.78, 12.21) | 8.38 (4.48, 12.28) |
|  | Black | 9.18 (4.89, 13.46) | 3.72 (-1.48, 8.92) | 4.06 (-4.24, 12.37) | 9.52 (5.93, 13.12) | 15.12 (10.36, 19.88) | 6.88 (2.91, 10.85) | 9.16 (5.06, 13.25) | 3.25 (-0.19, 6.69) | 19.41 (12.69, 26.14) |
| Education | Higher Education | 8.14 (6.73, 9.56) | 8.42 (6.69, 10.15) | 7.46 (5.78, 9.14) | 7.17 (5.58, 8.76) | 8.79 (6.90, 10.69) | 8.80 (6.67, 10.92) | 8.18 (6.48, 9.87) | 8.23 (6.67, 9.78) | 7.77 (5.64, 9.91) |
|  | Upper Secondary | 8.36 (6.73, 9.98) | 9.42 (7.64, 11.21) | 8.71 (6.52, 10.89) | 8.46 (6.47, 10.46) | 8.99 (7.04, 10.93) | 6.44 (4.59, 8.28) | 7.52 (5.69, 9.35) | 8.25 (6.20, 10.29) | 9.40 (7.15, 11.66) |
|  | Lower Secondary | 8.25 (6.84, 9.66) | 10.78 (9.08, 12.48) | 8.95 (7.21, 10.68) | 9.84 (7.85, 11.82) | 11.34 (9.43, 13.25) | 10.42 (8.63, 12.22) | 9.04 (7.24, 10.84) | 8.84 (6.97, 10.71) | 9.24 (7.11, 11.36) |
|  | No Qualifications | 9.16 (7.20, 11.12) | 9.68 (7.25, 12.12) | 10.70 (8.33, 13.08) | 10.08 (7.97, 12.19) | 8.78 (5.82, 11.75) | 9.20 (5.43, 12.97) | 11.52 (8.88, 14.17) | 7.72 (4.94, 10.50) | 11.22 (3.59, 18.86) |
| Financial stress | Low | 7.35 (6.11, 8.59) | 6.83 (5.52, 8.14) | 6.60 (5.16, 8.04) | 6.83 (5.62, 8.03) | 8.75 (7.27, 10.23) | 7.16 (5.78, 8.53) | 7.09 (5.82, 8.36) | 7.01 (5.67, 8.35) | 7.70 (6.15, 9.25) |
|  | Moderate | 7.08 (5.86, 8.31) | 9.11 (7.71, 10.51) | 8.45 (6.71, 10.19) | 8.65 (7.19, 10.11) | 8.34 (6.60, 10.09) | 8.26 (6.40, 10.11) | 7.76 (6.20, 9.32) | 8.88 (7.26, 10.50) | 7.82 (5.60, 10.04) |
|  | High | 7.52 (6.19, 8.84) | 9.44 (7.98, 10.90) | 8.32 (6.71, 9.94) | 8.13 (6.08, 10.18) | 9.70 (7.61, 11.79) | 8.23 (6.00, 10.45) | 8.18 (6.14, 10.22) | 5.35 (3.63, 7.07) | 6.43 (3.31, 9.55) |

**Supplementary Table 5.** Proportion of sample reporting a new diagnosis of depression across timepoints, excluding adults from future timepoints after first reporting a diagnosis of depression (%, 95% CI).

|  | Timepoint | 2011-2013 | 2012-2014 | 2013-2015 | 2014-2016 | 2015-2017 | 2016-2018 | 2017-2019 | 2019-2021 | 2020-2022 |
| --- | --- | --- | --- | --- | --- | --- | --- | --- | --- | --- |
| Caseness | No distress | 0.49 (0.40, 0.58) | 0.34 (0.26, 0.41) | 0.29 (0.22, 0.37) | 0.32 (0.24, 0.40) | 0.22 (0.15, 0.28) | 0.24 (0.17, 0.30) | 0.28 (0.20, 0.36) | 0.40 (0.28, 0.52) | 0.33 (0.23, 0.44) |
|  | Report distress | 4.15 (3.61, 4.69) | 3.23 (2.73, 3.73) | 2.78 (2.28, 3.28) | 2.67 (2.17, 3.18) | 2.88 (2.36, 3.41) | 2.41 (1.91, 2.91) | 2.70 (2.13, 3.27) | 3.37 (2.76, 3.98) | 2.06 (1.53, 2.60) |
| Sex | Men | 0.67 (0.55, 0.79) | 0.61 (0.47, 0.75) | 0.52 (0.38, 0.66) | 0.56 (0.41, 0.70) | 0.49 (0.36, 0.62) | 0.54 (0.39, 0.68) | 0.54 (0.39, 0.68) | 0.78 (0.58, 0.99) | 0.38 (0.19, 0.57) |
|  | Women | 1.56 (1.37, 1.75) | 1.07 (0.91, 1.22) | 0.96 (0.79, 1.12) | 0.79 (0.64, 0.95) | 0.81 (0.65, 0.96) | 0.66 (0.52, 0.80) | 0.88 (0.69, 1.08) | 1.14 (0.92, 1.36) | 0.99 (0.76, 1.22) |
| Age | 16-24 | 1.13 (0.79, 1.48) | 0.93 (0.65, 1.21) | 1.07 (0.69, 1.45) | 1.04 (0.70, 1.39) | 0.98 (0.65, 1.30) | 1.35 (0.93, 1.78) | 1.54 (1.00, 2.08) | 1.90 (1.29, 2.51) | 1.56 (0.82, 2.31) |
|  | 25-44 | 1.49 (1.25, 1.73) | 1.08 (0.86, 1.30) | 0.96 (0.72, 1.20) | 0.61 (0.42, 0.80) | 0.83 (0.62, 1.04) | 0.67 (0.46, 0.89) | 0.68 (0.46, 0.90) | 1.31 (0.93, 1.68) | 0.89 (0.55, 1.23) |
|  | 45-64 | 1.11 (0.92, 1.29) | 0.87 (0.69, 1.05) | 0.68 (0.50, 0.86) | 0.85 (0.65, 1.05) | 0.62 (0.45, 0.80) | 0.45 (0.30, 0.59) | 0.63 (0.44, 0.82) | 0.93 (0.69, 1.17) | 0.60 (0.42, 0.79) |
|  | 65+ | 0.64 (0.45, 0.82) | 0.44 (0.28, 0.59) | 0.35 (0.21, 0.50) | 0.30 (0.17, 0.43) | 0.29 (0.16, 0.43) | 0.29 (0.16, 0.41) | 0.39 (0.23, 0.55) | 0.21 (0.10, 0.32) | 0.20 (0.08, 0.33) |
| Age and sex | Male 16-24 | 0.62 (0.24, 0.99) | 0.43 (0.17, 0.70) | 0.49 (0.10, 0.89) | 0.68 (0.26, 1.10) | 0.51 (0.17, 0.85) | 1.19 (0.61, 1.78) | 1.20 (0.55, 1.85) | 1.21 (0.51, 1.91) | 1.16 (-0.06, 2.39) |
|  | Female 16-24 | 1.70 (1.13, 2.26) | 1.50 (0.98, 2.02) | 1.74 (1.07, 2.41) | 1.46 (0.86, 2.06) | 1.50 (0.91, 2.08) | 1.53 (0.91, 2.15) | 1.88 (1.02, 2.74) | 2.57 (1.59, 3.56) | 1.93 (1.04, 2.83) |
|  | Male 25-44 | 0.71 (0.47, 0.94) | 0.92 (0.60, 1.24) | 0.65 (0.35, 0.95) | 0.48 (0.21, 0.75) | 0.64 (0.37, 0.91) | 0.62 (0.31, 0.93) | 0.52 (0.26, 0.78) | 1.41 (0.83, 1.99) | 0.39 (0.15, 0.63) |
|  | Female 25-44 | 2.22 (1.81, 2.63) | 1.23 (0.93, 1.52) | 1.26 (0.92, 1.59) | 0.74 (0.48, 1.01) | 1.02 (0.69, 1.34) | 0.72 (0.45, 1.00) | 0.83 (0.50, 1.16) | 1.22 (0.75, 1.68) | 1.36 (0.77, 1.95) |
|  | Male 45-64 | 0.85 (0.61, 1.09) | 0.57 (0.34, 0.79) | 0.63 (0.38, 0.88) | 0.86 (0.56, 1.16) | 0.60 (0.33, 0.87) | 0.37 (0.18, 0.57) | 0.48 (0.24, 0.72) | 0.72 (0.39, 1.06) | 0.29 (0.12, 0.45) |
|  | Female 45-64 | 1.36 (1.07, 1.64) | 1.17 (0.89, 1.45) | 0.73 (0.47, 1.00) | 0.84 (0.58, 1.10) | 0.64 (0.42, 0.87) | 0.52 (0.31, 0.73) | 0.78 (0.48, 1.07) | 1.13 (0.78, 1.48) | 0.92 (0.59, 1.25) |
|  | Male 65+ | 0.39 (0.17, 0.60) | 0.39 (0.16, 0.61) | 0.23 (0.06, 0.41) | 0.15 (0.02, 0.28) | 0.16 (0.03, 0.29) | 0.25 (0.07, 0.43) | 0.23 (0.07, 0.40) | 0.05 (-0.02, 0.12) | 0.10 (-0.02, 0.22) |
|  | Female 65+ | 0.87 (0.57, 1.16) | 0.49 (0.27, 0.70) | 0.47 (0.23, 0.71) | 0.43 (0.21, 0.65) | 0.42 (0.18, 0.66) | 0.32 (0.15, 0.50) | 0.53 (0.27, 0.80) | 0.35 (0.15, 0.55) | 0.29 (0.08, 0.50) |
| Cohort | Pre-Boomers | 0.68 (0.49, 0.88) | 0.37 (0.22, 0.53) | 0.37 (0.19, 0.54) | 0.35 (0.18, 0.52) | 0.30 (0.14, 0.47) | 0.30 (0.13, 0.47) | 0.29 (0.11, 0.46) | 0.15 (-0.00, 0.31) | 0.24 (0.00, 0.47) |
|  | Baby Boomers | 1.02 (0.84, 1.20) | 0.86 (0.68, 1.05) | 0.60 (0.44, 0.76) | 0.71 (0.53, 0.90) | 0.47 (0.32, 0.62) | 0.40 (0.26, 0.53) | 0.52 (0.36, 0.68) | 0.48 (0.31, 0.64) | 0.38 (0.23, 0.53) |
|  | Generation X | 1.49 (1.23, 1.75) | 1.03 (0.80, 1.26) | 0.84 (0.63, 1.06) | 0.70 (0.48, 0.92) | 0.87 (0.63, 1.11) | 0.49 (0.32, 0.67) | 0.77 (0.52, 1.01) | 1.05 (0.75, 1.36) | 0.66 (0.44, 0.88) |
|  | Millennials | 1.25 (0.98, 1.52) | 1.02 (0.79, 1.25) | 1.11 (0.80, 1.42) | 0.87 (0.61, 1.12) | 0.79 (0.55, 1.03) | 1.05 (0.75, 1.35) | 1.19 (0.81, 1.56) | 1.67 (1.20, 2.15) | 1.04 (0.59, 1.49) |
| Ethnicity | White | 1.18 (1.05, 1.31) | 0.86 (0.75, 0.97) | 0.77 (0.65, 0.89) | 0.71 (0.59, 0.82) | 0.66 (0.55, 0.76) | 0.63 (0.52, 0.73) | 0.72 (0.60, 0.84) | 0.98 (0.81, 1.16) | 0.71 (0.55, 0.88) |
|  | Ethnic minorities | 0.55 (0.32, 0.77) | 0.67 (0.40, 0.94) | 0.48 (0.24, 0.72) | 0.32 (0.10, 0.54) | 0.59 (0.26, 0.91) | 0.26 (0.10, 0.42) | 0.64 (0.14, 1.14) | 0.82 (0.43, 1.22) | 0.53 (0.25, 0.80) |
|  | Mixed | 1.70 (0.58, 2.82) | 1.30 (0.15, 2.46) | 0.87 (0.03, 1.71) | 1.06 (-0.12, 2.24) | 0.22 (-0.01, 0.45) | 0.74 (-0.13, 1.62) | 1.98 (-0.66, 4.62) | 2.41 (0.41, 4.40) | 1.20 (-0.06, 2.46) |
|  | Asian | 0.31 (0.12, 0.51) | 0.51 (0.22, 0.80) | 0.32 (0.05, 0.59) | 0.24 (-0.02, 0.50) | 0.47 (0.13, 0.81) | 0.15 (0.03, 0.27) | 0.25 (0.06, 0.44) | 0.36 (0.07, 0.64) | 0.36 (0.11, 0.61) |
|  | Black | 0.58 (0.18, 0.97) | 0.67 (0.17, 1.16) | 0.50 (0.03, 0.98) | 0.19 (0.01, 0.38) | 0.39 (-0.15, 0.94) | 0.20 (-0.09, 0.48) | 0.26 (-0.14, 0.66) | 1.23 (0.25, 2.21) | 0.25 (-0.04, 0.53) |
| Education | Higher Education | 0.76 (0.60, 0.93) | 0.69 (0.53, 0.85) | 0.63 (0.47, 0.79) | 0.63 (0.46, 0.80) | 0.62 (0.45, 0.78) | 0.54 (0.39, 0.69) | 0.54 (0.38, 0.70) | 0.80 (0.59, 1.00) | 0.70 (0.50, 0.90) |
|  | Upper Secondary | 1.13 (0.87, 1.38) | 1.19 (0.92, 1.46) | 0.67 (0.44, 0.91) | 0.85 (0.59, 1.11) | 0.96 (0.70, 1.22) | 0.65 (0.43, 0.87) | 0.89 (0.59, 1.19) | 1.57 (1.11, 2.02) | 0.89 (0.55, 1.23) |
|  | Lower Secondary | 1.46 (1.18, 1.75) | 0.82 (0.61, 1.03) | 1.15 (0.84, 1.46) | 0.68 (0.44, 0.92) | 0.58 (0.36, 0.80) | 0.62 (0.40, 0.84) | 1.00 (0.65, 1.34) | 0.94 (0.62, 1.26) | 0.69 (0.35, 1.02) |
|  | No Qualifications | 1.33 (0.99, 1.67) | 0.71 (0.44, 0.98) | 0.69 (0.39, 0.98) | 0.73 (0.42, 1.04) | 0.49 (0.21, 0.77) | 0.56 (0.24, 0.89) | 0.63 (0.31, 0.94) | 0.53 (0.15, 0.90) | 0.26 (-0.01, 0.53) |
| Financial stress | Low | 0.58 (0.47, 0.69) | 0.46 (0.37, 0.55) | 0.48 (0.38, 0.58) | 0.47 (0.37, 0.57) | 0.47 (0.37, 0.57) | 0.43 (0.34, 0.52) | 0.41 (0.31, 0.52) | 0.56 (0.43, 0.69) | 0.49 (0.37, 0.60) |
|  | Moderate | 1.60 (1.33, 1.88) | 1.23 (0.97, 1.49) | 0.98 (0.68, 1.27) | 0.97 (0.69, 1.25) | 0.81 (0.56, 1.06) | 0.95 (0.66, 1.25) | 1.05 (0.75, 1.34) | 1.51 (1.07, 1.96) | 1.12 (0.66, 1.57) |
|  | High | 3.21 (2.58, 3.83) | 2.58 (2.00, 3.17) | 2.20 (1.59, 2.82) | 2.18 (1.44, 2.92) | 2.56 (1.64, 3.48) | 1.65 (0.91, 2.40) | 2.94 (1.89, 3.99) | 4.19 (2.68, 5.69) | 2.18 (0.68, 3.69) |

**Supplementary Table 6.** Mean GHQ-12 score for adults reporting a new diagnosis of depression across timepoints, excluding adults from future timepoints after first reporting a diagnosis of depression (mean, 95% CI).

|  | Timepoint | 2011-2013 | 2012-2014 | 2013-2015 | 2014-2016 | 2015-2017 | 2016-2018 | 2017-2019 | 2019-2021 | 2020-2022 |
| --- | --- | --- | --- | --- | --- | --- | --- | --- | --- | --- |
|  | Whole sample | 19.20 (18.27, 20.14) | 19.93 (18.87, 20.99) | 19.36 (18.05, 20.67) | 18.50 (17.29, 19.70) | 20.45 (19.13, 21.76) | 19.53 (18.18, 20.89) | 19.44 (18.19, 20.70) | 19.69 (18.56, 20.82) | 18.90 (17.33, 20.47) |
| Sex | Men | 18.37 (16.63, 20.12) | 18.81 (16.73, 20.89) | 18.99 (16.48, 21.51) | 17.44 (15.01, 19.88) | 21.43 (18.66, 24.19) | 20.05 (15.60, 24.51) | 19.83 (16.87, 22.78) | 18.99 (16.34, 21.65) | 19.69 (13.13, 26.25) |
|  | Women | 19.55 (18.44, 20.66) | 20.57 (19.23, 21.91) | 19.55 (17.94, 21.16) | 19.24 (17.71, 20.76) | 19.86 (18.19, 21.53) | 19.12 (17.38, 20.87) | 19.21 (17.74, 20.69) | 20.14 (18.62, 21.66) | 18.61 (16.60, 20.62) |
| Age | 16-24 | 18.80 (1.69, 35.91) | 19.89 (15.96, 23.82) | 19.41 | 20.32 (14.37, 26.26) | 21.23 (16.45, 26.01) | 18.97 (13.75, 24.18) | 19.41 | 19.55 (16.58, 22.52) | 18.58 (5.92, 31.24) |
|  | 25-44 | 19.49 (17.96, 21.02) | 20.24 (18.25, 22.22) | 19.59 (17.15, 22.04) | 17.05 (14.04, 20.07) | 20.39 (17.76, 23.02) | 21.36 (15.99, 26.73) | 17.79 (14.53, 21.05) | 17.91 (14.70, 21.12) | 19.03 (13.41, 24.66) |
|  | 45-64 | 19.47 (17.92, 21.02) | 20.58 (18.22, 22.94) | 20.17 (17.50, 22.84) | 19.06 (17.03, 21.08) | 20.38 (17.33, 23.43) | 16.95 (14.65, 19.25) | 20.52 (18.14, 22.90) | 21.56 (19.39, 23.74) | 18.87 (12.88, 24.86) |
|  | 65+ | 18.11 (15.56, 20.67) | 17.15 | 16.43 | 16.10 | 19.28 | 21.57 (14.57, 28.58) | 20.42 (5.63, 35.21) | 20.45 | 19.72 |
| Age and sex | Male 16-24 | 16.22 | 18.59 | 16.36 | 18.92 | 23.67 | 19.32 | 20.38 | 20.23 | 17.51 |
|  | Female 16-24 | 19.75 (1.14, 38.37) | 20.26 (14.78, 25.75) | 20.42 | 21.06 (0.83, 41.28) | 20.28 (0.20, 40.37) | 18.66 (10.95, 26.37) | 18.75 | 19.24 (14.75, 23.72) | 19.16 |
|  | Male 25-44 | 18.81 (15.32, 22.29) | 20.42 (15.10, 25.74) | 19.15 (-5.79, 44.09) | 15.05 | 22.57 (-1.11, 46.25) | 22.28 | 17.28 | 16.68 | 17.72 |
|  | Female 25-44 | 19.70 (17.81, 21.58) | 20.10 (15.11, 25.09) | 19.79 (16.93, 22.65) | 18.30 (14.85, 21.75) | 19.10 (13.33, 24.87) | 20.62 (14.15, 27.08) | 18.09 (1.53, 34.65) | 19.19 (13.65, 24.74) | 19.39 (-1.58, 40.36) |
|  | Male 45-64 | 19.62 (15.42, 23.82) | 18.86 | 20.36 (15.74, 24.99) | 18.66 (13.66, 23.66) | 20.29 (13.81, 26.77) | 16.92 | 21.08 (12.47, 29.69) | 21.47 (0.51, 42.44) | 21.50 |
|  | Female 45-64 | 19.37 (17.53, 21.21) | 21.37 (18.43, 24.31) | 20.01 | 19.48 (16.02, 22.93) | 20.46 (2.04, 38.89) | 16.97 (14.28, 19.66) | 20.16 (17.30, 23.01) | 21.62 (18.61, 24.64) | 18.04 |
|  | Male 65+ | 15.63 (-2.87, 34.13) | 13.85 | 17.21 | 12.73 | 17.10 | 22.36 | 20.84 | 16.30 | 33.35 |
|  | Female 65+ | 19.15 (3.12, 35.18) | 19.85 | 16.06 | 17.20 | 20.05 | 21.01 (11.10, 30.92) | 20.28 | 20.99 | 15.46 |
| Cohort | Pre-Boomers | 18.11 (15.56, 20.67) | 16.47 | 16.05 | 15.89 | 17.82 | 23.03 | 19.47 | 16.87 | 20.12 |
|  | Baby Boomers | 19.89 (18.14, 21.63) | 19.92 (17.51, 22.34) | 19.27 (15.84, 22.70) | 19.74 (17.37, 22.11) | 20.56 (16.59, 24.53) | 17.42 (14.25, 20.58) | 20.30 (17.63, 22.98) | 20.35 (15.43, 25.27) | 19.59 |
|  | Generation X | 19.78 (18.17, 21.38) | 20.43 (18.19, 22.66) | 19.81 (17.29, 22.33) | 17.60 (14.79, 20.41) | 21.38 (18.43, 24.33) | 19.61 (14.31, 24.92) | 19.48 (14.97, 23.99) | 22.18 (19.41, 24.94) | 19.18 (12.67, 25.68) |
|  | Millennials | 18.28 (15.50, 21.06) | 20.48 (17.85, 23.11) | 20.24 (16.57, 23.91) | 19.05 (15.78, 22.32) | 19.57 (14.42, 24.73) | 19.93 (16.08, 23.79) | 18.86 (4.34, 33.38) | 18.05 (14.80, 21.29) | 17.99 (11.86, 24.11) |
| Ethnicity | White | 19.22 (18.27, 20.17) | 20.00 (18.90, 21.10) | 19.20 (17.85, 20.56) | 18.40 (17.13, 19.67) | 20.01 (18.61, 21.41) | 19.59 (18.17, 21.00) | 19.50 (18.17, 20.82) | 19.74 (18.54, 20.95) | 18.77 (17.01, 20.52) |
|  | Ethnic minorities | 17.05 (12.58, 21.53) | 19.54 (-8.04, 47.12) | 21.57 (-10.06, 53.19) | 19.74 (14.20, 25.28) | 25.79 (20.76, 30.82) | 21.71 (17.68, 25.73) | 18.87 (13.66, 24.08) | 19.06 (-0.99, 39.10) | 20.43 (14.12, 26.74) |
|  | Mixed | 16.03 | 22.70 | 25.10 | 17.84 (-22.73, 58.42) | 23.35 | 19.60 | 23.64 | 22.71 | 21.13 |
|  | Asian | 14.38 (5.42, 23.34) | 19.58 (-27.46, 66.62) | 26.02 | 21.84 (-2.00, 45.67) | 22.79 (15.94, 29.63) | 25.87 (17.01, 34.73) | 14.88 (5.75, 24.01) | 18.63 | 20.28 (-8.78, 49.34) |
|  | Black | 21.76 | 16.10 | 12.54 | 19.52 | 25.65 | 17.30 (-14.81, 49.41) | 15.00 | 14.65 | 18.28 |
| Education | Higher Education | 18.70 (15.99, 21.41) | 18.73 (15.41, 22.06) | 17.97 (15.02, 20.92) | 16.61 (4.90, 28.33) | 18.86 (16.08, 21.65) | 19.51 (16.10, 22.93) | 19.30 (16.05, 22.56) | 19.86 (17.42, 22.30) | 17.71 (12.66, 22.75) |
|  | Upper Secondary | 19.50 (17.23, 21.77) | 20.14 (15.64, 24.65) | 18.02 (-2.83, 38.87) | 18.74 (2.47, 35.01) | 20.52 (5.76, 35.29) | 16.98 (2.49, 31.47) | 19.33 (3.94, 34.73) | 18.63 (5.05, 32.21) | 20.21 (14.55, 25.86) |
|  | Lower Secondary | 19.30 (17.37, 21.22) | 21.25 (18.66, 23.84) | 20.22 (17.54, 22.90) | 21.07 (17.06, 25.07) | 21.71 (5.44, 37.97) | 21.92 (18.48, 25.36) | 19.20 (15.39, 23.01) | 20.61 (17.29, 23.92) | 20.65 (0.61, 40.69) |
|  | No Qualifications | 19.52 (17.18, 21.85) | 18.94 (-0.87, 38.76) | 22.42 (17.35, 27.48) | 19.20 (13.70, 24.69) | 18.81 (12.63, 24.99) | 18.56 (9.50, 27.63) | 21.11 (15.78, 26.43) | 17.69 | 10.75 |
| Financial stress | Low | 16.48 (14.94, 18.03) | 16.69 (14.70, 18.67) | 16.69 (14.52, 18.86) | 16.08 (14.32, 17.84) | 18.83 (16.70, 20.97) | 17.75 (15.97, 19.53) | 17.29 (15.34, 19.24) | 17.41 (15.57, 19.24) | 17.86 (15.19, 20.52) |
|  | Moderate | 19.07 (17.43, 20.71) | 20.63 (18.89, 22.36) | 20.57 (16.94, 24.19) | 20.38 (18.17, 22.60) | 20.13 (15.37, 24.90) | 21.04 (17.44, 24.64) | 18.62 (16.47, 20.78) | 21.25 (18.39, 24.11) | 19.38 (15.46, 23.30) |
|  | High | 22.63 (21.04, 24.23) | 23.11 (20.40, 25.82) | 22.65 (18.95, 26.36) | 21.90 (17.51, 26.29) | 25.06 (21.48, 28.65) | 22.69 (16.11, 29.28) | 23.68 (17.63, 29.73) | 21.49 (18.77, 24.22) | 21.52 (11.98, 31.06) |

**Supplementary Table 7.** Proportion of sample reporting psychological distress without a recent depression diagnosis across timepoints, excluding adults from future timepoints after first reporting a diagnosis of depression (%, 95% CI).

|  | Timepoint | 2011-2013 | 2012-2014 | 2013-2015 | 2014-2016 | 2015-2017 | 2016-2018 | 2017-2019 | 2019-2021 | 2020-2022 |
| --- | --- | --- | --- | --- | --- | --- | --- | --- | --- | --- |
|  | Whole sample | 16.52 (16.07, 16.97) | 16.13 (15.67, 16.59) | 17.24 (16.76, 17.72) | 14.70 (14.25, 15.16) | 15.43 (14.95, 15.91) | 16.46 (15.95, 16.97) | 16.92 (16.36, 17.49) | 18.89 (18.24, 19.53) | 20.85 (20.17, 21.53) |
| Sex | Men | 14.11 (13.49, 14.72) | 13.19 (12.60, 13.79) | 14.87 (14.22, 15.52) | 12.32 (11.71, 12.92) | 12.93 (12.29, 13.58) | 13.90 (13.21, 14.59) | 13.77 (13.02, 14.53) | 15.55 (14.72, 16.38) | 16.84 (15.91, 17.77) |
|  | Women | 18.85 (18.25, 19.45) | 18.98 (18.34, 19.62) | 19.56 (18.90, 20.21) | 17.05 (16.40, 17.69) | 17.87 (17.21, 18.52) | 18.93 (18.22, 19.65) | 19.91 (19.14, 20.69) | 22.03 (21.14, 22.92) | 24.63 (23.70, 25.56) |
| Age | 16-24 | 19.61 (18.36, 20.86) | 19.29 (17.93, 20.64) | 19.19 (17.93, 20.44) | 17.51 (16.20, 18.82) | 19.00 (17.58, 20.42) | 19.22 (17.71, 20.74) | 22.60 (20.87, 24.33) | 23.83 (21.92, 25.74) | 27.05 (24.98, 29.12) |
|  | 25-44 | 17.39 (16.60, 18.17) | 17.42 (16.58, 18.27) | 17.98 (17.10, 18.86) | 15.78 (14.90, 16.66) | 16.70 (15.76, 17.63) | 17.52 (16.46, 18.58) | 19.26 (18.10, 20.42) | 22.15 (20.78, 23.52) | 24.82 (23.38, 26.26) |
|  | 45-64 | 17.11 (16.37, 17.85) | 16.58 (15.81, 17.36) | 18.41 (17.57, 19.24) | 14.75 (14.02, 15.48) | 15.47 (14.68, 16.26) | 16.41 (15.57, 17.25) | 16.77 (15.89, 17.66) | 19.00 (18.01, 19.98) | 19.63 (18.58, 20.67) |
|  | 65+ | 12.46 (11.58, 13.33) | 11.91 (11.08, 12.73) | 13.55 (12.61, 14.49) | 11.62 (10.80, 12.45) | 11.90 (11.06, 12.74) | 13.80 (12.93, 14.67) | 11.50 (10.65, 12.35) | 13.18 (12.18, 14.17) | 15.76 (14.68, 16.84) |
| Age and sex | Male 16-24 | 15.46 (13.82, 17.11) | 14.68 (13.00, 16.36) | 15.61 (13.93, 17.28) | 13.37 (11.68, 15.06) | 14.47 (12.64, 16.29) | 15.07 (13.12, 17.01) | 16.66 (14.44, 18.87) | 18.15 (15.59, 20.71) | 19.78 (16.93, 22.63) |
|  | Female 16-24 | 24.09 (22.29, 25.90) | 24.53 (22.57, 26.48) | 23.31 (21.35, 25.26) | 22.24 (20.28, 24.21) | 24.09 (21.96, 26.22) | 23.83 (21.61, 26.06) | 28.43 (25.94, 30.92) | 29.18 (26.53, 31.83) | 33.60 (30.67, 36.53) |
|  | Male 25-44 | 15.16 (14.06, 16.27) | 14.68 (13.49, 15.87) | 16.76 (15.46, 18.06) | 13.92 (12.68, 15.16) | 14.61 (13.27, 15.95) | 15.35 (13.93, 16.76) | 16.65 (15.04, 18.26) | 19.24 (17.37, 21.12) | 21.65 (19.51, 23.79) |
|  | Female 25-44 | 19.44 (18.38, 20.51) | 20.00 (18.86, 21.14) | 19.13 (17.98, 20.29) | 17.57 (16.38, 18.76) | 18.67 (17.39, 19.95) | 19.57 (18.11, 21.02) | 21.71 (20.10, 23.32) | 24.82 (22.96, 26.69) | 27.83 (25.88, 29.78) |
|  | Male 45-64 | 14.72 (13.69, 15.74) | 14.18 (13.12, 15.24) | 15.50 (14.40, 16.61) | 12.67 (11.68, 13.66) | 13.10 (12.04, 14.17) | 13.67 (12.51, 14.83) | 13.70 (12.50, 14.90) | 15.77 (14.43, 17.11) | 16.03 (14.57, 17.50) |
|  | Female 45-64 | 19.42 (18.41, 20.43) | 18.88 (17.81, 19.94) | 21.19 (20.04, 22.34) | 16.78 (15.75, 17.82) | 17.78 (16.72, 18.85) | 19.07 (17.92, 20.23) | 19.79 (18.53, 21.04) | 22.19 (20.79, 23.59) | 23.22 (21.74, 24.70) |
|  | Male 65+ | 10.81 (9.59, 12.02) | 8.90 (7.88, 9.93) | 11.21 (9.98, 12.44) | 9.15 (8.03, 10.27) | 9.82 (8.69, 10.94) | 11.87 (10.68, 13.06) | 9.15 (8.06, 10.24) | 10.47 (9.28, 11.66) | 11.98 (10.60, 13.35) |
|  | Female 65+ | 14.02 (12.81, 15.22) | 14.73 (13.49, 15.97) | 15.78 (14.50, 17.06) | 13.92 (12.74, 15.11) | 13.82 (12.66, 14.99) | 15.56 (14.31, 16.80) | 13.66 (12.43, 14.90) | 15.66 (14.24, 17.07) | 19.15 (17.65, 20.65) |
| Cohort | Pre-Boomers | 12.55 (11.63, 13.47) | 12.08 (11.18, 12.98) | 13.83 (12.76, 14.91) | 11.96 (10.98, 12.95) | 12.51 (11.45, 13.57) | 14.81 (13.62, 16.01) | 12.17 (10.96, 13.38) | 13.32 (11.88, 14.76) | 15.99 (14.39, 17.59) |
|  | Baby Boomers | 16.82 (16.05, 17.58) | 15.87 (15.08, 16.65) | 17.27 (16.42, 18.12) | 14.18 (13.42, 14.93) | 14.29 (13.50, 15.08) | 14.52 (13.72, 15.32) | 14.10 (13.23, 14.97) | 16.56 (15.57, 17.55) | 17.40 (16.36, 18.44) |
|  | Generation X | 16.83 (16.03, 17.64) | 16.77 (15.89, 17.66) | 18.30 (17.38, 19.22) | 14.76 (13.93, 15.58) | 15.62 (14.72, 16.52) | 17.25 (16.26, 18.23) | 17.82 (16.75, 18.89) | 19.38 (18.21, 20.55) | 20.75 (19.52, 21.99) |
|  | Millennials | 19.36 (18.35, 20.37) | 19.06 (18.03, 20.08) | 18.59 (17.57, 19.61) | 16.86 (15.77, 17.95) | 17.80 (16.65, 18.96) | 18.31 (17.06, 19.56) | 20.09 (18.68, 21.51) | 22.88 (21.28, 24.48) | 25.64 (23.96, 27.32) |
| Ethnicity | White | 16.17 (15.70, 16.64) | 15.89 (15.41, 16.37) | 16.97 (16.45, 17.48) | 14.52 (14.04, 15.01) | 15.37 (14.86, 15.88) | 16.35 (15.81, 16.89) | 16.76 (16.17, 17.36) | 18.71 (18.03, 19.39) | 20.75 (20.02, 21.48) |
|  | Ethnic minorities | 20.61 (19.17, 22.05) | 18.82 (17.40, 20.24) | 20.29 (18.74, 21.83) | 16.61 (15.18, 18.03) | 16.24 (14.90, 17.58) | 17.85 (16.27, 19.42) | 18.43 (16.63, 20.24) | 20.59 (18.62, 22.57) | 21.78 (19.69, 23.87) |
|  | Mixed | 20.00 (16.00, 24.00) | 17.27 (13.25, 21.28) | 22.67 (18.17, 27.17) | 20.40 (16.29, 24.50) | 21.46 (17.03, 25.89) | 22.93 (18.11, 27.75) | 25.37 (20.05, 30.69) | 25.77 (20.40, 31.14) | 24.86 (19.37, 30.35) |
|  | Asian | 20.77 (18.80, 22.73) | 18.63 (16.77, 20.48) | 20.84 (18.71, 22.96) | 14.84 (13.25, 16.43) | 15.16 (13.44, 16.88) | 15.47 (13.56, 17.38) | 16.52 (14.49, 18.54) | 19.68 (17.14, 22.23) | 20.80 (18.13, 23.46) |
|  | Black | 19.28 (16.85, 21.71) | 19.39 (16.68, 22.11) | 17.37 (14.64, 20.10) | 16.08 (13.80, 18.35) | 16.43 (13.89, 18.96) | 18.25 (15.27, 21.23) | 17.88 (14.35, 21.41) | 20.82 (16.95, 24.69) | 21.86 (17.38, 26.35) |
| Education | Higher Education | 15.86 (15.12, 16.59) | 15.36 (14.64, 16.09) | 16.27 (15.52, 17.02) | 14.57 (13.82, 15.31) | 14.74 (13.99, 15.48) | 15.30 (14.50, 16.09) | 16.07 (15.18, 16.97) | 19.92 (18.94, 20.90) | 21.91 (20.88, 22.93) |
|  | Upper Secondary | 16.47 (15.55, 17.40) | 16.19 (15.23, 17.16) | 17.41 (16.42, 18.39) | 14.69 (13.70, 15.69) | 15.77 (14.71, 16.83) | 16.99 (15.89, 18.09) | 18.34 (17.19, 19.50) | 19.26 (17.94, 20.57) | 21.79 (20.34, 23.25) |
|  | Lower Secondary | 16.99 (16.07, 17.91) | 17.17 (16.20, 18.15) | 17.86 (16.82, 18.89) | 14.54 (13.56, 15.52) | 16.00 (14.93, 17.07) | 17.18 (16.06, 18.31) | 17.44 (16.23, 18.66) | 18.07 (16.77, 19.38) | 19.96 (18.52, 21.40) |
|  | No Qualifications | 17.27 (16.02, 18.52) | 16.58 (15.27, 17.88) | 18.37 (16.97, 19.77) | 16.18 (14.78, 17.57) | 17.88 (16.33, 19.42) | 18.06 (16.48, 19.63) | 17.56 (15.79, 19.34) | 16.79 (14.72, 18.87) | 18.64 (16.37, 20.91) |
| Financial stress | Low | 11.75 (11.27, 12.24) | 11.76 (11.28, 12.24) | 12.87 (12.36, 13.38) | 11.41 (10.94, 11.89) | 12.13 (11.65, 12.61) | 12.74 (12.23, 13.25) | 12.46 (11.90, 13.03) | 14.84 (14.19, 15.49) | 16.61 (15.94, 17.28) |
|  | Moderate | 20.35 (19.43, 21.28) | 19.49 (18.49, 20.49) | 21.60 (20.53, 22.66) | 19.28 (18.18, 20.38) | 21.13 (19.91, 22.35) | 22.63 (21.32, 23.93) | 24.46 (23.05, 25.87) | 25.39 (23.85, 26.93) | 29.60 (27.82, 31.38) |
|  | High | 36.84 (35.10, 38.58) | 38.50 (36.57, 40.43) | 41.30 (39.11, 43.48) | 40.10 (37.50, 42.70) | 41.15 (38.27, 44.03) | 44.09 (41.06, 47.12) | 43.85 (41.02, 46.68) | 46.90 (43.63, 50.17) | 51.28 (47.29, 55.27) |

**Supplementary Table 8.** Mean difference in psychological distress (GHQ-12 score) between adults with a new diagnosis of depression and adults without a new diagnosis of depression across timepoints, excluding adults from future timepoints after first reporting a diagnosis of depression (*b*, 95% CI).

|  | Timepoint | 2011-2013 | 2012-2014 | 2013-2015 | 2014-2016 | 2015-2017 | 2016-2018 | 2017-2019 | 2019-2021 | 2020-2022 |
| --- | --- | --- | --- | --- | --- | --- | --- | --- | --- | --- |
|  | Overall sample | 8.56 (7.64, 9.48) | 9.36 (8.34, 10.39) | 8.59 (7.34, 9.84) | 8.15 (7.00, 9.30) | 9.96 (8.68, 11.23) | 8.81 (7.50, 10.12) | 8.51 (7.30, 9.73) | 8.39 (7.27, 9.51) | 7.34 (5.91, 8.77) |
| Sex | Men | 8.24 (6.60, 9.88) | 8.83 (7.11, 10.56) | 8.73 (6.60, 10.86) | 7.54 (5.68, 9.39) | 11.41 (9.09, 13.72) | 9.80 (7.76, 11.83) | 9.39 (7.12, 11.66) | 8.22 (6.19, 10.25) | 8.81 (5.81, 11.80) |
|  | Women | 8.41 (7.32, 9.49) | 9.44 (8.19, 10.68) | 8.29 (6.82, 9.76) | 8.45 (7.01, 9.89) | 8.92 (7.39, 10.44) | 7.95 (6.30, 9.60) | 7.82 (6.46, 9.17) | 8.34 (6.90, 9.78) | 6.41 (4.74, 8.09) |
| Age | 16-24 | 8.20 (5.55, 10.85) | 9.49 (6.71, 12.28) | 8.90 (6.10, 11.71) | 9.92 (7.21, 12.63) | 10.69 (8.03, 13.36) | 8.12 (5.74, 10.51) | 7.92 (5.22, 10.62) | 7.61 (5.34, 9.88) | 6.18 (4.18, 8.18) |
|  | 25-44 | 8.64 (7.21, 10.08) | 9.38 (7.73, 11.03) | 8.66 (6.53, 10.78) | 6.52 (4.21, 8.82) | 9.64 (7.46, 11.82) | 10.45 (8.02, 12.89) | 6.39 (4.37, 8.41) | 6.02 (4.03, 8.00) | 6.73 (4.16, 9.30) |
|  | 45-64 | 8.56 (7.11, 10.00) | 9.75 (8.09, 11.41) | 9.01 (6.94, 11.07) | 8.58 (6.80, 10.35) | 9.68 (7.20, 12.16) | 6.01 (4.05, 7.97) | 9.46 (7.34, 11.57) | 10.15 (8.25, 12.05) | 7.35 (4.61, 10.09) |
|  | 65+ | 8.13 (6.04, 10.22) | 7.25 (4.71, 9.80) | 6.24 (3.54, 8.93) | 6.18 (3.98, 8.38) | 9.40 (6.37, 12.43) | 11.40 (8.20, 14.59) | 10.47 (8.17, 12.77) | 10.16 (4.76, 15.56) | 9.19 (1.20, 17.17) |
| Age and sex | Male 16-24 | 6.43 (1.35, 11.51) | 9.04 (5.37, 12.72) | 6.67 (0.62, 12.73) | 9.32 (4.45, 14.20) | 13.94 (9.00, 18.88) | 9.35 (6.22, 12.48) | 9.87 (4.83, 14.91) | 9.21 (5.33, 13.10) | 6.50 (2.84, 10.17) |
|  | Female 16-24 | 8.26 (5.38, 11.14) | 8.88 (5.49, 12.27) | 8.96 (6.13, 11.79) | 9.74 (6.62, 12.86) | 8.84 (5.70, 11.97) | 6.85 (3.32, 10.38) | 6.29 (3.40, 9.18) | 6.43 (3.66, 9.19) | 5.49 (3.36, 7.62) |
|  | Male 25-44 | 8.64 (7.21, 10.08) | 10.06 (7.63, 12.50) | 8.53 (4.67, 12.39) | 4.80 (0.54, 9.05) | 12.17 (8.51, 15.84) | 11.65 (7.53, 15.77) | 6.22 (2.92, 9.53) | 5.28 (2.44, 8.12) | 5.96 (2.66, 9.25) |
|  | Female 25-44 | 8.38 (6.69, 10.07) | 8.76 (6.48, 11.04) | 8.55 (6.17, 10.92) | 7.50 (5.05, 9.94) | 8.02 (5.39, 10.64) | 9.45 (6.54, 12.35) | 6.37 (3.80, 8.94) | 6.84 (4.30, 9.38) | 6.57 (3.32, 9.82) |
|  | Male 45-64 | 8.56 (7.11, 10.00) | 8.52 (5.02, 12.02) | 9.71 (6.75, 12.68) | 8.61 (6.34, 10.87) | 10.04 (5.92, 14.16) | 6.47 (2.71, 10.23) | 10.47 (6.51, 14.42) | 10.55 (7.28, 13.83) | 10.48 (4.46, 16.50) |
|  | Female 45-64 | 8.03 (6.39, 9.66) | 10.05 (8.24, 11.86) | 8.35 (5.43, 11.27) | 8.59 (5.84, 11.33) | 9.32 (6.47, 12.17) | 5.56 (3.50, 7.61) | 8.65 (6.32, 10.99) | 9.72 (7.40, 12.03) | 6.02 (3.14, 8.89) |
|  | Male 65+ | 8.13 (6.04, 10.22) | 4.55 (0.80, 8.31) | 7.50 (4.69, 10.30) | 3.26 (1.06, 5.45) | 7.64 (1.88, 13.39) | 12.59 (8.34, 16.83) | 11.35 (6.49, 16.20) | 6.42 (-0.25, 13.10) | 23.50 (20.43, 26.56) |
|  | Female 65+ | 8.76 (6.21, 11.32) | 9.39 (6.51, 12.26) | 5.41 (1.71, 9.10) | 6.86 (4.20, 9.52) | 9.77 (6.20, 13.35) | 10.47 (5.95, 14.98) | 9.91 (7.29, 12.53) | 10.32 (4.29, 16.36) | 4.31 (-3.81, 12.42) |
| Cohort | Pre-Boomers | 8.11 (6.02, 10.20) | 6.55 (3.72, 9.38) | 5.82 (2.70, 8.94) | 5.93 (3.72, 8.13) | 7.88 (4.41, 11.35) | 12.75 (8.74, 16.76) | 9.41 (6.81, 12.01) | 6.60 (-5.16, 18.37) | 9.63 (-3.57, 22.84) |
|  | Baby Boomers | 9.04 (7.48, 10.60) | 9.22 (7.51, 10.92) | 8.32 (5.90, 10.74) | 9.40 (7.33, 11.47) | 10.11 (7.60, 12.61) | 6.87 (4.63, 9.11) | 9.79 (7.57, 12.01) | 9.45 (7.21, 11.70) | 8.60 (5.42, 11.78) |
|  | Generation X | 8.89 (7.37, 10.40) | 9.62 (7.72, 11.51) | 8.73 (6.60, 10.86) | 7.08 (4.82, 9.33) | 10.66 (8.30, 13.02) | 8.58 (6.13, 11.02) | 8.21 (6.15, 10.28) | 10.62 (8.39, 12.85) | 7.43 (4.45, 10.41) |
|  | Millennials | 7.61 (5.67, 9.55) | 9.83 (7.82, 11.84) | 9.57 (7.31, 11.84) | 8.59 (6.28, 10.90) | 8.90 (6.54, 11.26) | 9.03 (6.63, 11.43) | 7.35 (5.10, 9.60) | 6.06 (4.04, 8.07) | 5.57 (2.77, 8.38) |
| Ethnicity | White | 8.60 (7.67, 9.54) | 9.44 (8.38, 10.51) | 8.45 (7.16, 9.74) | 8.04 (6.84, 9.23) | 9.52 (8.20, 10.83) | 8.86 (7.50, 10.21) | 8.57 (7.31, 9.84) | 8.47 (7.28, 9.65) | 7.24 (5.68, 8.79) |
|  | Ethnic minorities | 6.13 (2.90, 9.36) | 8.85 (4.60, 13.11) | 10.59 (5.69, 15.48) | 9.52 (6.11, 12.94) | 15.37 (11.55, 19.20) | 11.02 (7.96, 14.09) | 7.97 (3.63, 12.31) | 7.52 (4.41, 10.63) | 8.64 (5.75, 11.53) |
|  | Mixed | 5.19 (-0.16, 10.53) | 12.20 (4.10, 20.29) | 13.70 (10.13, 17.27) | 6.97 (0.67, 13.27) | 11.82 (4.08, 19.55) | 7.79 (1.93, 13.66) | 11.73 (9.87, 13.59) | 10.61 (6.97, 14.25) | 8.57 (2.34, 14.81) |
|  | Asian | 3.35 (-0.75, 7.46) | 8.69 (1.41, 15.96) | 14.93 (8.62, 21.23) | 11.74 (8.05, 15.43) | 12.43 (7.59, 17.27) | 15.40 (11.34, 19.46) | 4.08 (-0.12, 8.28) | 7.02 (1.40, 12.64) | 8.50 (4.00, 12.99) |
|  | Black | 11.23 (6.60, 15.87) | 5.85 (-1.44, 13.14) | 2.13 (-7.82, 12.08) | 9.74 (6.16, 13.32) | 15.85 (10.73, 20.96) | 7.12 (2.14, 12.11) | 4.66 (4.12, 5.20) | 3.52 (-0.52, 7.57) | 7.01 (2.96, 11.05) |
| Education | Higher Education | 8.23 (6.55, 9.90) | 8.33 (6.28, 10.37) | 7.42 (5.34, 9.49) | 6.39 (4.58, 8.21) | 8.52 (6.23, 10.81) | 8.97 (6.56, 11.39) | 8.52 (6.23, 10.82) | 8.57 (6.61, 10.53) | 6.04 (3.73, 8.36) |
|  | Upper Secondary | 8.93 (6.99, 10.86) | 9.66 (7.62, 11.71) | 7.31 (4.09, 10.53) | 8.46 (5.96, 10.97) | 10.02 (7.74, 12.31) | 6.24 (3.99, 8.48) | 8.21 (5.83, 10.59) | 7.18 (5.08, 9.29) | 8.48 (5.90, 11.07) |
|  | Lower Secondary | 8.53 (6.76, 10.31) | 10.52 (8.47, 12.57) | 9.30 (7.26, 11.34) | 10.56 (8.10, 13.03) | 11.05 (8.54, 13.56) | 11.04 (8.61, 13.48) | 8.12 (5.77, 10.47) | 9.24 (6.67, 11.80) | 9.09 (5.98, 12.21) |
|  | No Qualifications | 8.62 (6.38, 10.86) | 8.18 (5.12, 11.25) | 11.29 (8.16, 14.42) | 8.43 (5.91, 10.95) | 7.95 (5.11, 10.79) | 7.56 (3.42, 11.70) | 10.14 (6.32, 13.95) | 6.63 (3.32, 9.95) | -0.34 (-6.02, 5.33) |
| Financial stress | Low | 6.72 (5.34, 8.11) | 6.97 (5.33, 8.61) | 6.71 (4.85, 8.58) | 6.35 (4.83, 7.88) | 8.98 (7.18, 10.78) | 7.72 (6.13, 9.31) | 7.18 (5.61, 8.76) | 6.88 (5.24, 8.52) | 7.07 (5.17, 8.97) |
|  | Moderate | 7.62 (6.06, 9.18) | 9.25 (7.72, 10.77) | 8.88 (6.64, 11.12) | 8.98 (7.22, 10.74) | 8.43 (6.26, 10.60) | 9.01 (6.47, 11.54) | 6.20 (4.34, 8.06) | 8.61 (6.58, 10.65) | 6.10 (3.31, 8.90) |
|  | High | 8.38 (6.87, 9.88) | 8.59 (6.65, 10.53) | 7.79 (5.47, 10.12) | 7.30 (4.56, 10.03) | 9.98 (7.41, 12.55) | 7.24 (4.20, 10.28) | 8.05 (5.24, 10.86) | 4.95 (2.95, 6.96) | 4.76 (0.39, 9.12) |

